# 3D genomic features across >50 diverse cell types reveal insights into the genomic architecture of childhood obesity

**DOI:** 10.1101/2023.08.30.23294092

**Authors:** Khanh B. Trang, Matthew C. Pahl, James A. Pippin, Chun Su, Sheridan H. Littleton, Prabhat Sharma, Nikhil N. Kulkarni, Louis R. Ghanem, Natalie A. Terry, Joan M. O’Brien, Yadav Wagley, Kurt D. Hankenson, Ashley Jermusyk, Jason W. Hoskins, Laufey T. Amundadottir, Mai Xu, Kevin M Brown, Stewart A. Anderson, Wenli Yang, Paul M. Titchenell, Patrick Seale, Laura Cook, Megan K. Levings, Babette S. Zemel, Alessandra Chesi, Andrew D. Wells, Struan F.A. Grant

## Abstract

The prevalence of childhood obesity is increasing worldwide, along with the associated common comorbidities of type 2 diabetes and cardiovascular disease in later life. Motivated by evidence for a strong genetic component, our prior genome-wide association study (GWAS) efforts for childhood obesity revealed 19 independent signals for the trait; however, the mechanism of action of these loci remains to be elucidated. To molecularly characterize these childhood obesity loci we sought to determine the underlying causal variants and the corresponding effector genes within diverse cellular contexts. Integrating childhood obesity GWAS summary statistics with our existing 3D genomic datasets for 57 human cell types, consisting of high-resolution promoter-focused Capture-C/Hi-C, ATAC-seq, and RNA-seq, we applied stratified LD score regression and calculated the proportion of genome-wide SNP heritability attributable to cell type-specific features, revealing pancreatic alpha cell enrichment as the most statistically significant. Subsequent chromatin contact-based fine-mapping was carried out for genome-wide significant childhood obesity loci and their linkage disequilibrium proxies to implicate effector genes, yielded the most abundant number of candidate variants and target genes at the *BDNF*, *ADCY3*, *TMEM18* and *FTO* loci in skeletal muscle myotubes and the pancreatic beta-cell line, EndoC-BH1. One novel implicated effector gene, *ALKAL2* – an inflammation-responsive gene in nerve nociceptors – was observed at the key TMEM18 locus across multiple immune cell types. Interestingly, this observation was also supported through colocalization analysis using expression quantitative trait loci (eQTL) derived from the Genotype-Tissue Expression (GTEx) dataset, supporting an inflammatory and neurologic component to the pathogenesis of childhood obesity. Our comprehensive appraisal of 3D genomic datasets generated in a myriad of different cell types provides genomic insights into pediatric obesity pathogenesis.

**KEY POINTS:** *Question:* What are the causal variants and corresponding effector genes conferring pediatric obesity susceptibility in different cellular contexts?

*Findings:* Our method of assessing 3D genomic data across a range of cell types revealed heritability enrichment of childhood obesity variants, particularly within pancreatic alpha cells. The mapping of putative causal variants to cis-regulatory elements revealed candidate effector genes for cell types spanning metabolic, neural, and immune systems.

*Meaning:* We gain a systemic view of childhood obesity genomics by leveraging 3D techniques that implicate regulatory regions harboring causal variants, providing insights into the disease pathogenesis across different cellular systems.

## INTRODUCTION

The prevalence of obesity has risen significantly worldwide^1^, especially among children and adolescents^2^. Obesity is associated with chronic diseases, such as diabetes, cardiovascular diseases, and certain cancers^3–6^, along with mechanical issues including osteoarthritis and sleep apnea^7^.

Modern lifestyle factors, including physical inactivity, excessive caloric intake, and socioeconomic inequity, along with disrupted sleep and microbiome, represent environmental risk factors for obesity pathogenesis. However, genetics also play a significant role, with the estimated heritability ranging from 40% to 70%^8–10^. Studies show that body weight and obesity remain stable from infancy to adulthood^11–14^, but variation between individuals does exist^15^. Genome-wide association studies (GWAS) have improved our understanding of the genetic contribution to childhood obesity^16–21^. However, the functional consequences and molecular mechanisms of identified genetic variants in such GWAS efforts are yet to be fully elucidated. Efforts are now being made to predict target effector genes and explore potential drug targets using various computational and experimental approaches^22–26^, which subsequently warrant functional follow-up efforts.

With our extensive datasets generated on a range of different cell types, by combining 3D chromatin maps (Hi-C, Capture-C) with matched transcriptome (RNA-seq) and chromatin accessibility data (ATAC-seq), we investigated heritability patterns of pediatric obesity-associated variants and their gene-regulatory functions in a cell type-specific manner. This approach yielded 94 candidate causal variants mapped to their putative effector gene(s) and corresponding cell type(s) setting. In addition, using methods comparable to our prior efforts in other disease contexts^27–34^, we also uncovered new variant-to-gene combinations within specific novel cellular settings, most notably in immune cell types, which further confirmed the involvement of the immune system in the pathogenesis of obesity in the early stages of life.

## METHODS

### Data and resource

Datasets used in prior studies are listed in **eTable 1**. ATAC-seq, RNA-seq, Hi-C, and Capture-C *library generation* for each cell type is provided in their original published study and their *pre-processing pipelines and tools* can be found in **eMethods**.

### Definition of cis-Regulatory Elements (cREs)

We intersected ATAC-seq open chromatin regions (OCRs) of each cell type with chromatin conformation capture data determined by Hi-C/Capture-C of the same cell type, and with promoters (-1,500/+500bp of TSS, which were referenced by GENCODE v30.

### Childhood obesity GWAS summary statistics

Data on childhood obesity from the EGG consortium was downloaded from www.egg-consortium.org. We used 8,566,179 European ancestry variants (consisting of 8,613 cases and 12,696 controls in stage I; of 921 cases and 1,930 controls in stage II), representing ∼55% of the total 15,504,218 variants observed across all ancestries in the original study^35^. The sumstats file was reformatted by *munge_sumstats.py* to standardize with the weighted variants from HapMap v3 within the LDSC baseline, which reduced the variants to 1,217,311 (7.8% of total).

### Cell type specific partitioned heritability

We used *LDSC* (http://www.github.com/bulik/ldsc) *v.1.0.1* with *--h2* flag to estimate SNP-based heritability of childhood obesity within 4 defined sets of input genomic regions: (1) OCRs, (2) OCRs at gene promoters, (3) cREs, and (4) cREs with an expanded window of ±500 bp. The baseline model LD scores, plink filesets, allele frequencies and variants weights files for the European 1000 genomes project phase 3 in hg38 were downloaded from the provided link (https://alkesgroup.broadinstitute.org/LDSCORE/GRCh38/). The cREs of each cell type were used to create the annotation, which in turn were used to compute annotation-specific LD scores for each cell types cREs set.

### Genetic loci included in variant-to-genes mapping

19 sentinel signals that achieved genome-wide significance in the trans-ancestral meta-analysis study^35^ were leveraged for our analyses. Proxies for each sentinel SNP were queried using TopLD^36^ and LDlinkR tool^37^ with the GRCh38 Genome assembly, 1000 Genomes phase 3 v5 variant set, European population, and LD threshold of r^2^>0.8, which resulted in 771 proxies, including the 21 SNPs from the 99% credible set of the original study (**eTable 2**).

### GWAS-eQTL colocalization

The summary statistics for the European ancestry subset from the EGG consortium GWAS for childhood obesity was used. Common variants (MAF□≥□0.01) from the 1000 Genomes Project v3 samples were used as a reference panel. We used non-overlapped genomic windows of ±250,000 bases extended in both directions from the median genomic position of each of 19 sentinel loci as input. We used ColocQuiaL^38^ to test genome-wide colocalization of all possible variants included in each inputted window against GTEx v.8 eQTLs associations for all 49 tissues available from https://www.gtexportal.org/home/datasets. A conditional posterior probability of colocalization of 0.8 or greater was imposed.

## RESULTS

### Enrichment assessment of childhood obesity variants across cell types

To explore the enrichment of childhood obesity GWAS variants across cell types, we carried out Partitioned Linkage Disequilibrium Score Regression (LDSR)^39^ on all ATAC-seq-defined OCRs for each cell type. We assessed cell-type specific enrichment of GWAS signals in four main categories of genomic regions (**Fig. 1A**): (1) Total OCRs: open chromatin regions defined by ATAC-seq; (2) Promoter OCRs: the subset of OCRs overlapping a gene promoter; (3) cREs: the subset of OCRs that form chromatin loops (as determined by Hi-C/Promoter Capture-C) with a gene promoter, and are therefore considered putative enhancers or suppressors regulating gene expression; (4) cREs ± 500bases: extended cREs by 500 bases in both directions. The rationale behind this approach is that different GWAS variants can influence phenotypes by regulating gene expression in a cell-type specific manner through various regulatory mechanisms. For example, they may alter enhancer function (cREs category) or affect the binding of a transcription factor at a gene’s promoter (Promoter OCRs category).

**Figure 1:**
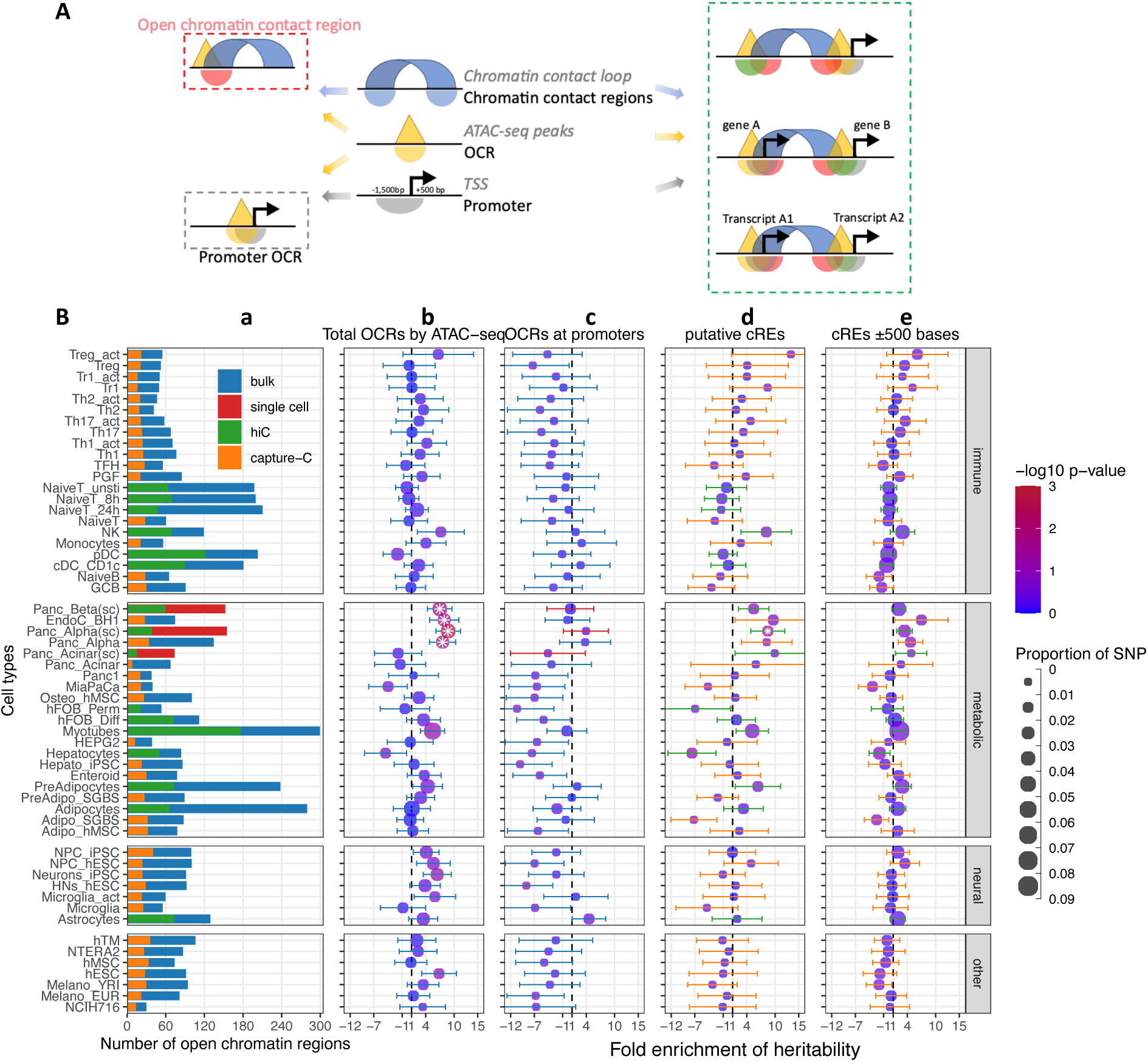
Partitioned Linkage Disequilibrium Score Regression analysis for open chromatin regions of all cell types. A. The schematic shows the different types of regions defined in our study and 3 different ways overlapping chromatin contact regions – OCRs – gene promoters define cREs. B. Heritability enrichment by LDSC analysis for each cell type

a) Bar-plot shows the total number of OCRs identified by ATAC-seq for each cell type on bulk cells - blue, or on single cells – red; and the portion of OCRs that fall within cREs identified by incorporating Hi-C (green) or by Capture-C (orange).
b-e) 4 panels of dot-plots show heritability enrichment by LDSC analysis for each cell type, with standard error whiskers. Dots’ colors correspond to -log10(p-values), dots with white asterisks are significant p-values <0.05, and dots’ sizes corresponding to the proportion of SNP contribute to heritability. Dash line at 1, i.e., no enrichment.

b) Analysis done on whole OCRs set of each cell type (whiskers colors match with bulk/single cell from bar-plot a);
c) On only OCRs that overlapped with promoters (whiskers’ colors match with bulk/single cell from bar-plot a);
d) On the putative cREs of each cell type (whiskers’ colors match with Hi-C/Capture-C from bar-plot a);
e) On the same cREs as (c) panel with their genomic positions expanded ±500 bases on both sides (whiskers’ colors match with Hi-C/Capture-C from bar-plot a)

We observed that 41 of 57 cell types – including 22 metabolic, 21 immune, 7 neural cell types and 7 independent cell lines (**eTable 1**) – showed at least a degree of directional enrichment with the total set of OCRs (**Fig. 1B – Total OCRs**). However, only four cell types – two pancreatic alpha and two pancreatic beta cell-based datasets – had statistically significant enrichments (*P*<0.05). These enrichments were less pronounced when focusing on promoter OCRs only (**Fig. 1B – Promoter OCRs**). To further limit the LD enrichment assessment to just those OCRs that can putatively regulate gene expression via chromatin contacts with gene promoters, we used the putative cREs^27,29^. This reduced the number of cell types showing at least nominal enrichment (31 of 57), enlarged the dispersion of enrichment ranges across different cell types, increased the 95% confidence intervals (CI) of enrichments, and hence increased the *P*-value of the resulting regression score. cREs from pancreatic alpha cells derived from single-cell ATAC-seq were the only dataset that remained statically significant (**Fig. 1B – putative cREs**).

The original reported LDSR method analyzed enrichment in the 500bp flanking regions of their regulatory categories^39^. However, when we expanded our analysis to the ±500bp window for our cREs, albeit incorporating more weighted variants into the enrichment (represented by larger dots in **Fig. 1B – cREs ±500 bases**), this resulted in a decrease in the number of cell types yielding at least nominal enrichment (26 cell types), the enrichment range across cell types, the 95% CI, and level of significance. The pancreatic alpha cell observation also dropped below the bar for significance with this expanded window definition.

### Consistency and diversity of childhood obesity proxy variants mapped to cREs

Despite the enrichments above only being limited to just a small number of cell types, it is likely that individual loci have differing levels of contributions in various cellular contexts and could not be detected at the genome wide assessment scale. As such we elected to further explore the candidate effector genes that are directly affected by cREs harboring childhood obesity-associated variants by systematically mapping the genomic positions of the LD proxies onto each cell type’s cREs. Most proxies fall within chromatin contact regions (**blue area in Venn diagram Fig. 2A**) or OCRs (**yellow area**) or open chromatin contact regions (**red area**), or completely outside (**white area**) any defined region. Only 94 proxies fall within our defined cREs (**overlapped area with dotted green border in Fig. 2A**), they clustered at 13 original loci (**eTable 3**). **eFig. 1A** outlines the number of signals at each locus included or excluded based on the criteria we defined for our regions of interest. The *TMEM18* locus yielded the most variants through cREs mapping, with 46 proxies for the two lead independent variants, rs7579427 and rs62104180. The second most abundant locus was *ADCY3*, with 21 proxies for lead variant rs4077678 (**Fig. 2B**). The higher number of variants at one locus did not correlate with implicating more genes or cell types through mapping. The mapping frequency of various variants within a specific locus exhibited substantial differences.

**Figure 2:**
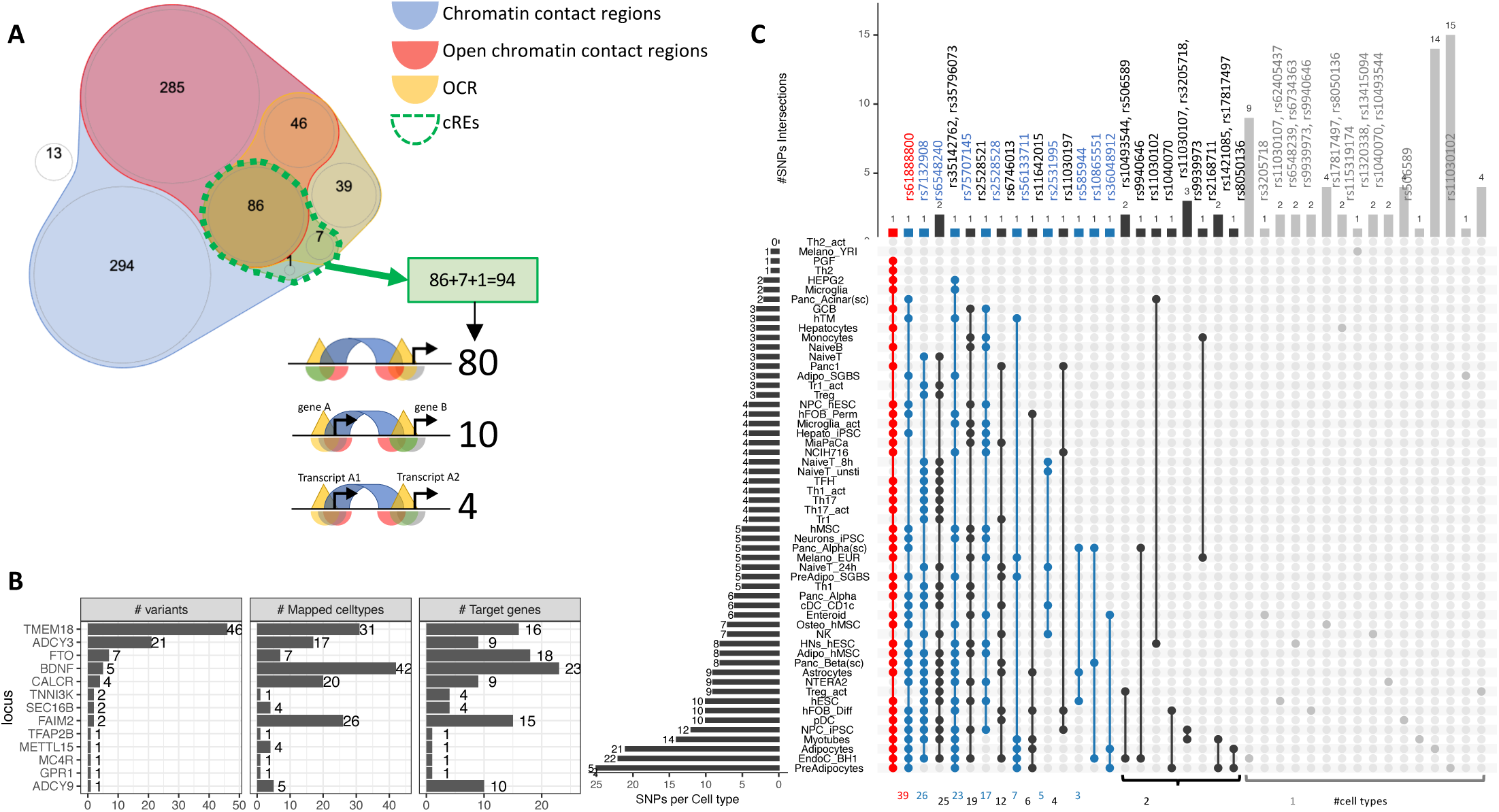
Mapping 771 proxies to the open chromatin regions of each cell type. A. Venn diagram shows how 771 proxies mapped to the OCRs:

- Blue area: 758 proxies were located within contact regions of at least one cell type regardless of chromatin state;
- Red area: 417 proxies were located within contact regions marked as open by overlapping with OCR;
- Yellow area: If we only considered open chromatin regions, 178 proxies were included;
- Dotted green bordered area: To focus on just those variants residing within open chromatin and contacting promoter regions in any cell type, we overlapped the genomic positions of these proxies with each cell type’s cRE set, yielding 90 variants (3 from the 99% credible set) directly contacting open gene promoters **(eTable 3)**, with 10 of which located within a promoter of one gene but contacting another different gene promoter. There were an additional 4 variants located within gene promoters but in chromatin contact with promoter(s) of nearby transcript(s) of the same gene (correspond to 3 cREs illustrations in Figure 1A).
- White area: proxies that fall into neither defined region of interest. B. Bar-plot shows number of proxies, cell types and target genes mapped at each locus. C. The upSet plot shows the degree of overlap across cell types of the variants; ranked from the most common variant (red) – rs61888800 from *BDNF* locus, a well-known 5’ untranslated region variant of this gene that is associated with anti-depression and therapeutic response^81,82^ – appeared in 39 cell types, to the group of variants (grey) which appeared in only one cell type.

Inspecting individual variants regardless of their locus, we found that 28 of 94 proxies appeared in cREs across multiple cell types, with another 66 observed in just one cell type (**Fig. 2D)**. 45 variants of these 66 just contacted one gene promoter, such as at the *GPR1* and *TFAP2B* loci (**eFig. 2**).

Overall, the number of cell types in which a variant was observed in open chromatin correlated with the number of genes contacted via chromatin loops (**eFig. 3A**). However, we also observed that some variants found in cREs in multiple cell types were more selective with respect to their candidate effector genes (**eFig. 3B-red arrow**), or conversely, more selective across given cell types but implicated multiple genes (**eFig. 3B-blue arrow**). **eFig. 4** outlines our observations at the *TMEM18* locus – an example locus involved in both scenarios.

**Figure 3:**
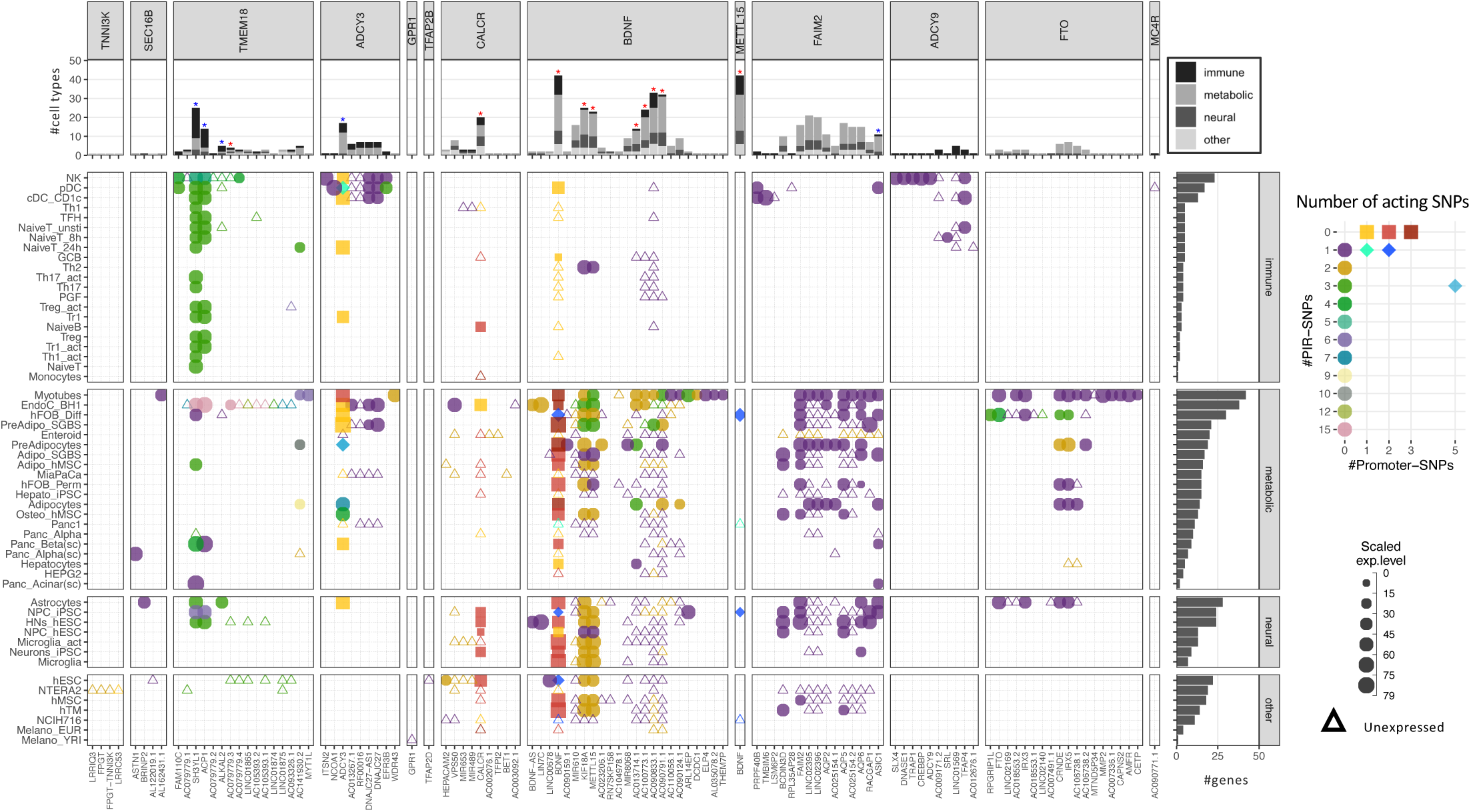
Profiles of 111 implicated genes by 94 proxies through cREs of each cell type. Main panel: Bubble plot show corresponding expression level (size) and number of variants (color) target each implicated gene of each cell type. Squares represent genes with variants at their promoters. Circles represent genes with variants contacted through chromatin loops. Some genes were implicated by both types, these “double implications” are represented as diamond shapes, and were identified across several cell types: two cell types (plasmacytoid dendritic cells and pre-differentiated adipocytes) for *ADCY3* gene, and five for *BDNF* (human embryonic stem cells - hESC, differentiated human fetal osteoblast cells - hFOB_Diff, neural progenitor cells derived from induced pluripotent stem cells - NPC_iPSC, PANC-1, and NCIH716 cell lines)
Genes with expression undetected in our arrays are shown as triangles. Top panel: bar-plot shows numbers of cell types each gene was implicated within, color-coded by which systems the cell types belong to. Right panel: bar-plot shows numbers of genes implicated by the variants with each cell type.

**Figure 4:**
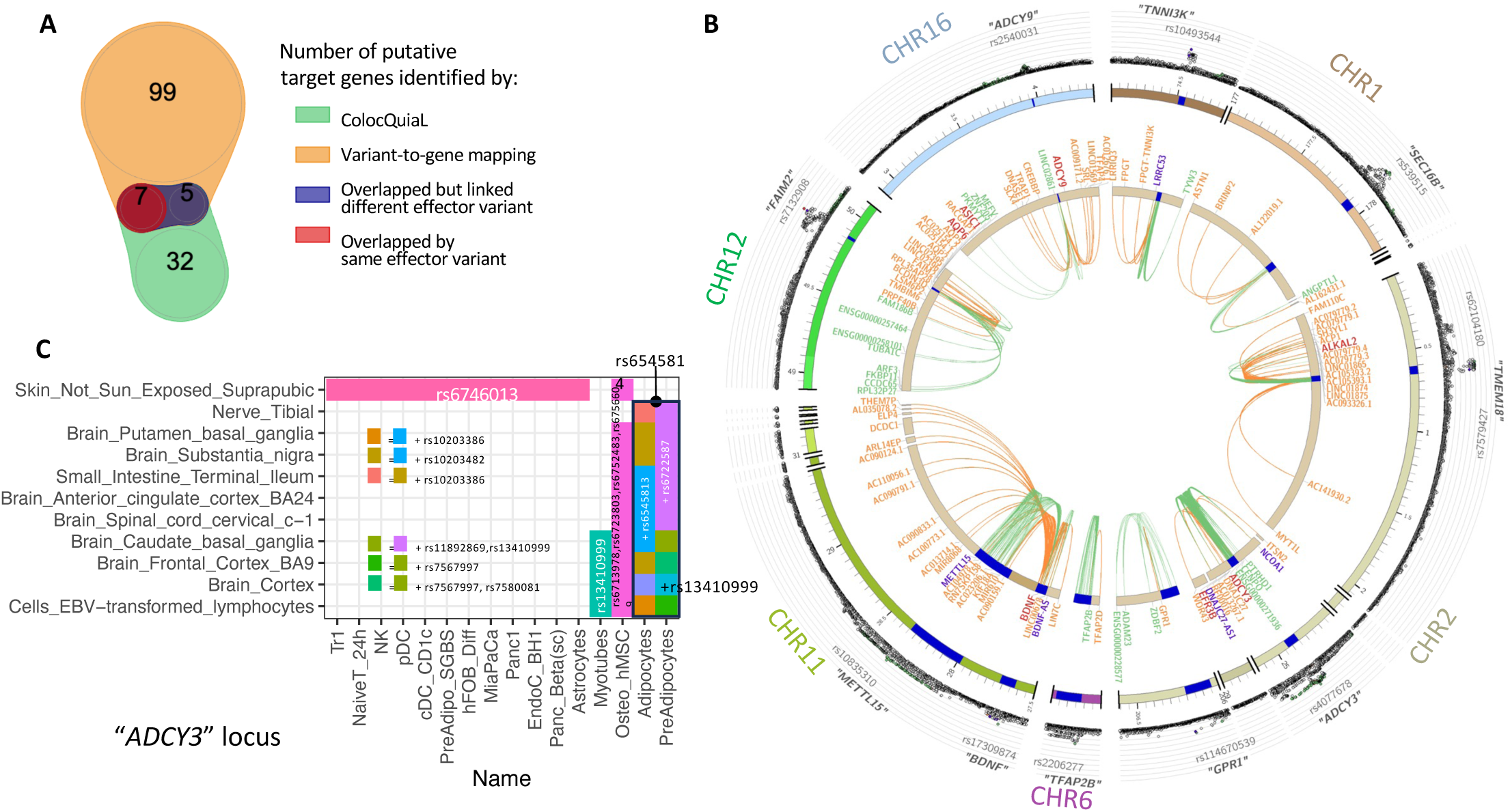
Colocalization of target effector genes with eQTLs. A. Venn diagram shows the overlaps between sets of genes yielded by ColocQuiaL and the variant-to-gene mapping process. B. Circos plot of the 10 loci demonstrates the differences in the ranges of associations between the two approaches, with long-ranged chromatin contacts between obesity variants and target genes displayed as orange links and short-range eQTLs colocalizations as green links.

Two SNPs – rs35796073, and rs35142762 within the *TMEM18* locus, in linkage disequilibrium with rs7579427 – were estimated with high probability (cond.PP.H4=0.78) of colocalizing with the expression of *ALKAL2* gene in subcutaneous adipose tissue. These pairs of SNP-gene were also identified by our variant-to-gene mapping approach in natural killer cells, plasmacytoid dendritic cells, unstimulated PBMC naïve CD4 T cells and astrocytes.
The rs7132908 variant at the *FAIM2* locus colocalized with the expression of *AQP6* in thyroid tissue and with *ASIC1* in prostate tissue, not only with high cond.PP.H4 but also with high individual SNP causal probability (SNP.PP.H4 > 0.95). rs7132908 was the second most consistent observation in our variant-to-gene mapping, namely across 25 different cell types (Figure 2B) and all three systems plus the other independent cell lines. The pair of rs7132908-contacting-*AQP6* was observed in 15 different cell types - 8 metabolic and 4 neural cell types, and 3 independent cell lines. The pair of rs7132908-contacting-*ASIC1* was observed in 11 different cell types - 8 metabolic and 2 neural cell types, and plasmacytoid dendritic cells.
The other eQTL signals that overlapped with our variant-to-gene mapping results were: *BDNF* at the *METTL15* locus with its promoter physically contacted by rs11030197 in 4 cell types and its expression significantly colocalized (cond.PP.H4=0.82) in tibial artery; *ADCY9* at its locus with its promoter physically contacted by rs2531995 in natural killer cells and its expression significantly colocalized in skin tissue (“Skin_Not_Sun_Exposed_Suprapubic”, cond.PP.H4=0.97). And *ADCY3* in the C panel. C. ColocQuiaL estimated that these SNPs highly colocalize with the expression of *ADCY3* in 11 different tissues, where the overlapping with the 16 cell types is represented, color-coded by the proxies rs numbers.

### Implicated genes cluster at loci strongly associated with childhood obesity consistently across multiple cell types

Mapping the variants across all the cell types resulted in a total of 111 implicated childhood obesity candidate effector genes (**Table 1**). Among these, 45 genes were specific to just one cell type (**eFig. 5A**), including 13 in myotubes and 7 in natural killer cells. Conversely and notably, *BDNF* appeared across 42 different cell types. Across the metabolic, neural, and immune systems and seven other cell lines, there were 9 genes consistently implicated in all four categories (top panel **Fig. 3 –** red stars, **eFig. 5B**: “all”), while 5 genes were consistently implicated in metabolic, neural, and immune systems (top panel **Fig. 3** – blue stars, **eFig. 5B**: “all_main”). Two genes, *ADCY3* and *BDNF*, had variants both at their promoters and contacted variants in cREs via chromatin loops (**eFig. 6**).

**Table 1.**
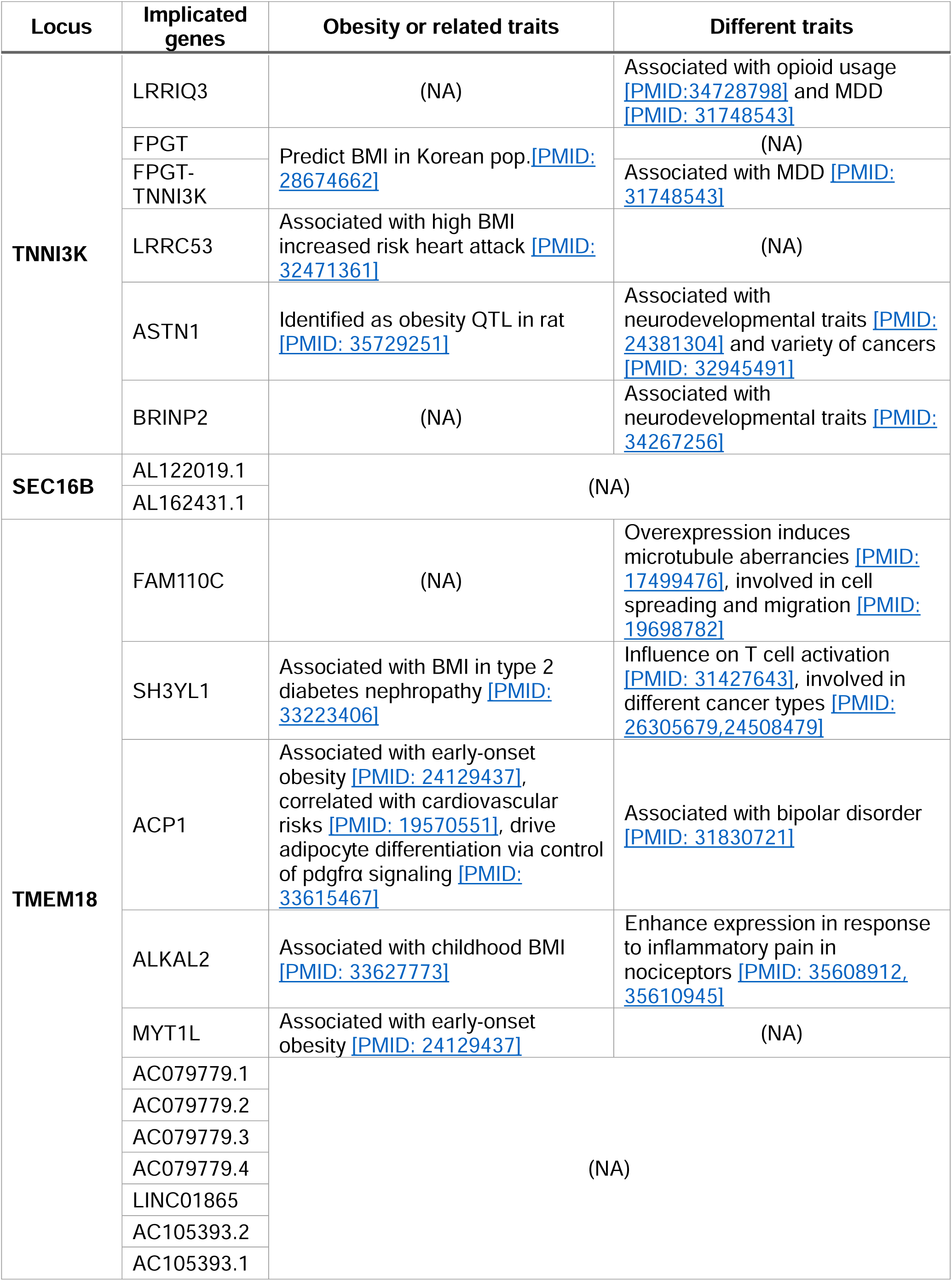
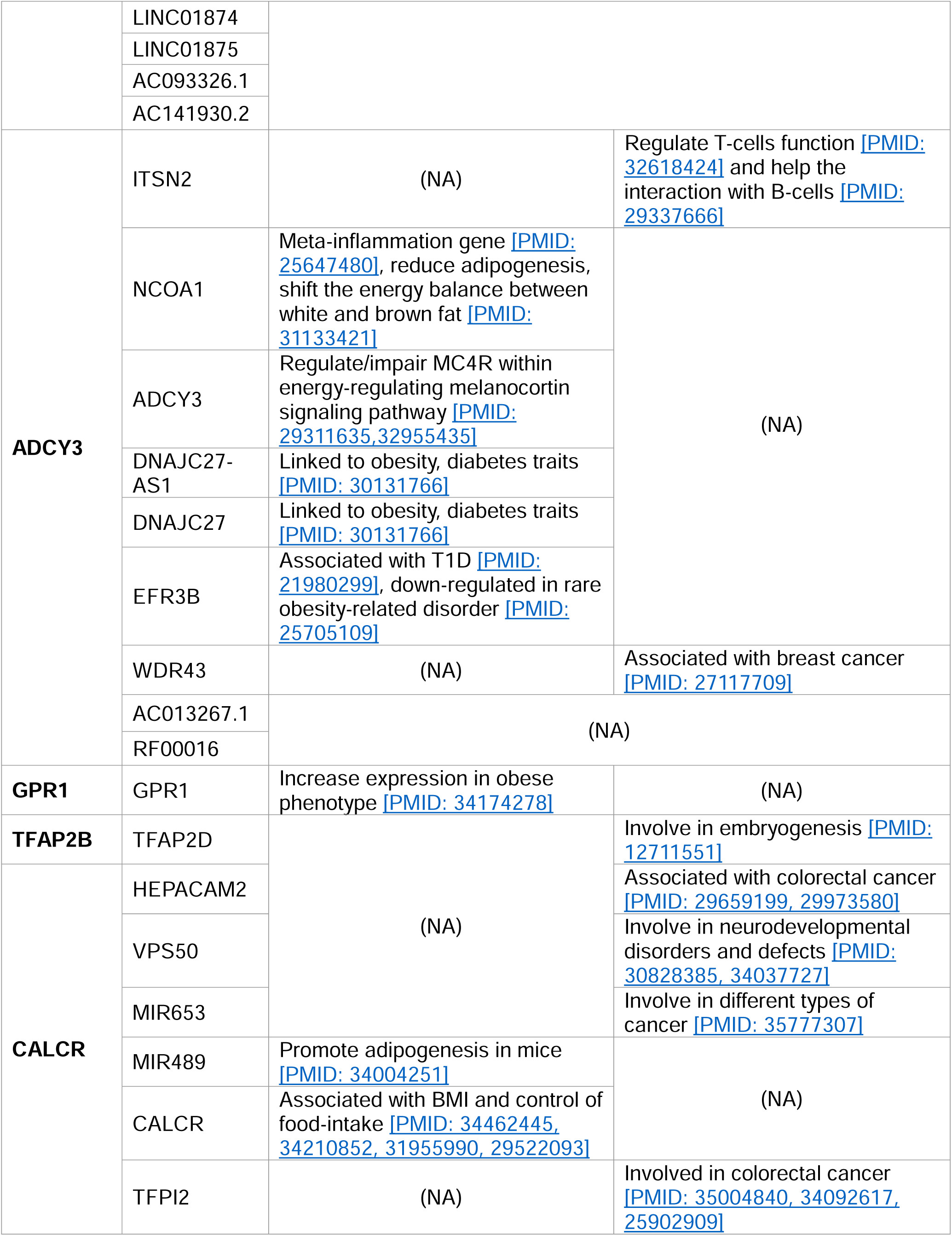
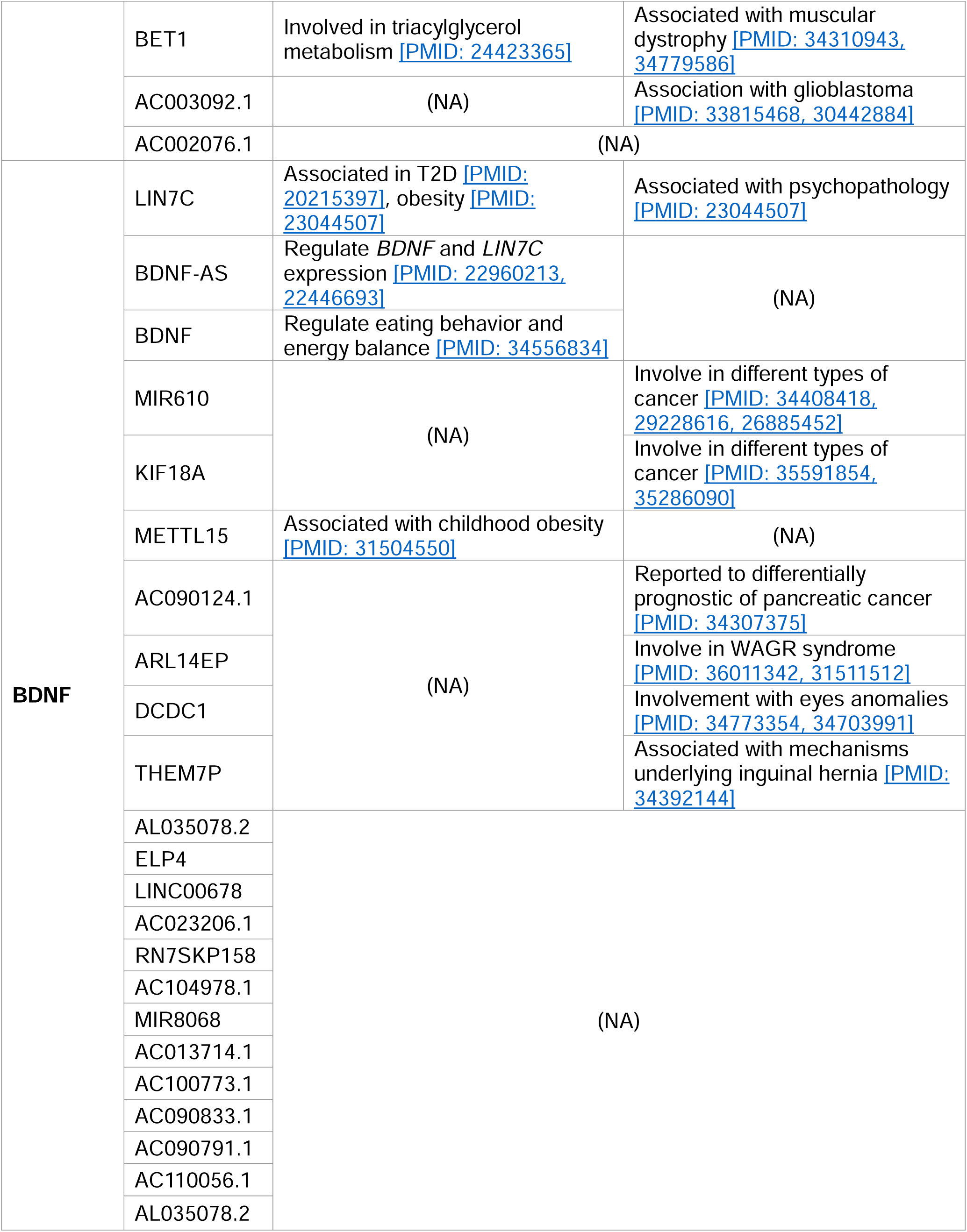
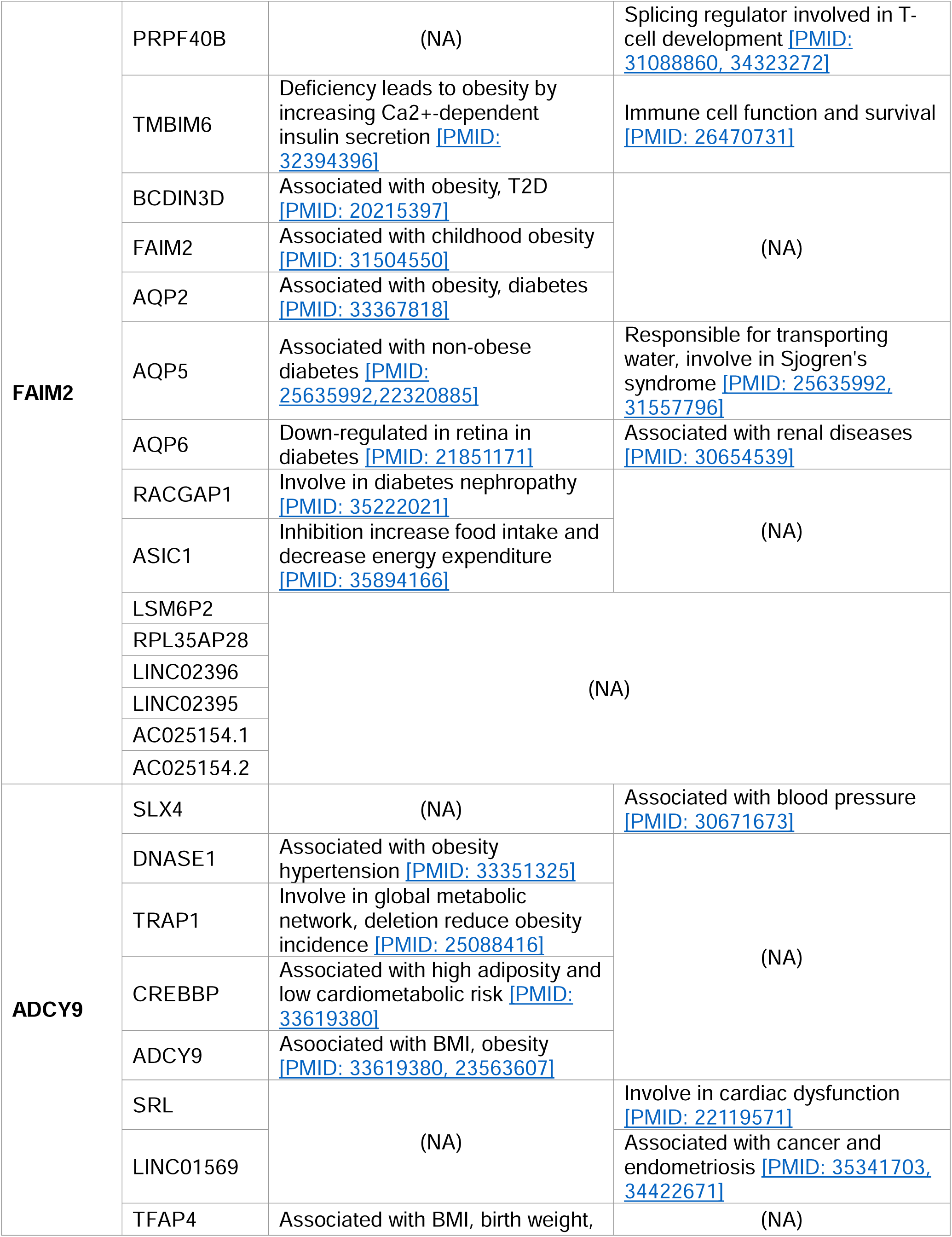
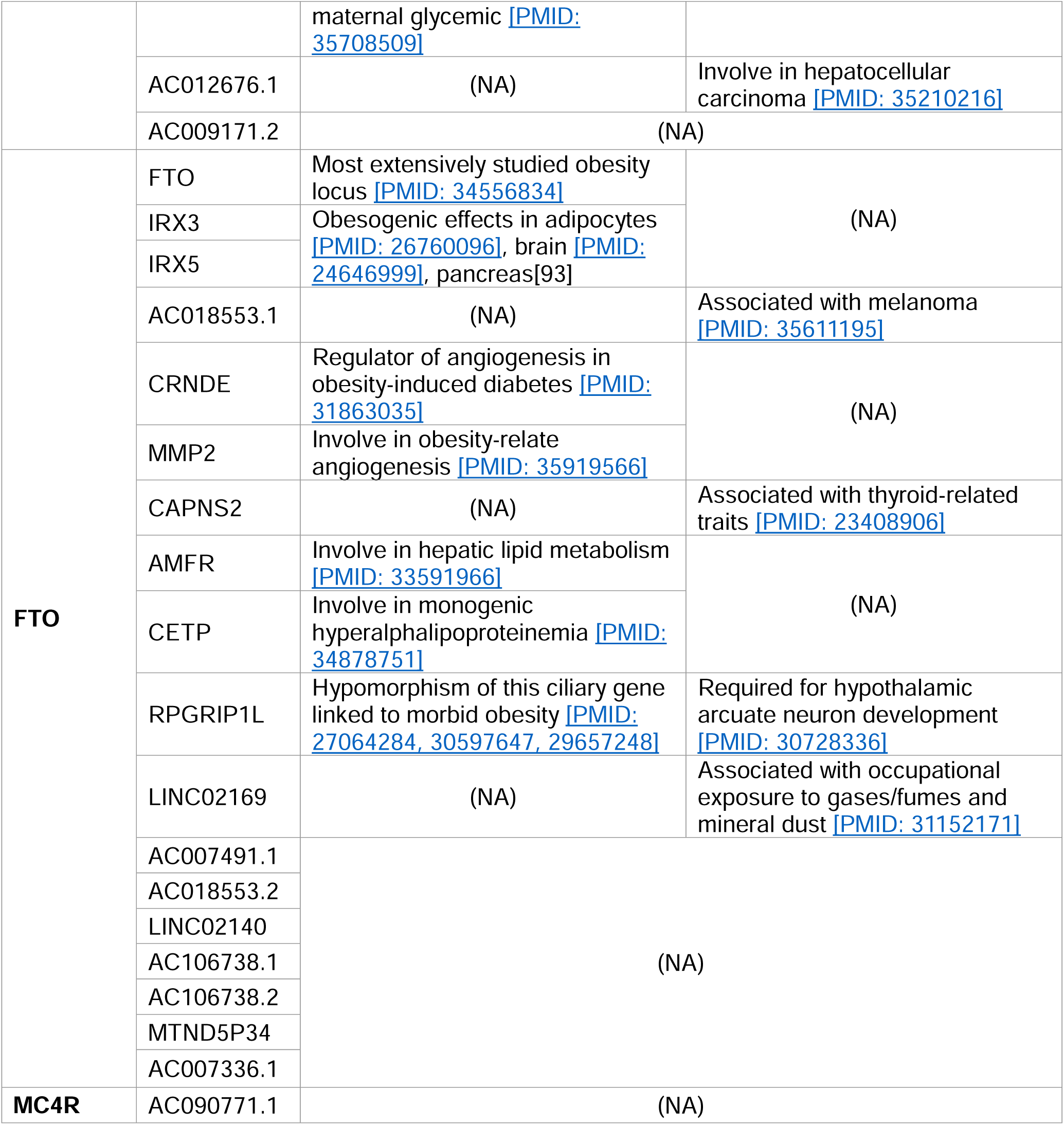
PubMed-query known functions for 111 genes implicated by obesity variants.

At the *TMEM18* locus on chr 2p25.3, a highly significant human obesity locus that has long been associated with both adult and childhood obesity, we obeserved differing degrees of evidence for 16 genes, but noted that rs6548240, rs35796073, and rs35142762 consistently contacted the *SH3YL1, ACP1*, and *ALKAL2* promoters across multiple cell types (**Fig. 2D-third and fourth column**).

At the chr 2p23 locus, *ADCY3* yielded the most contacts (i.e. many proxies contacting the same gene via chromatin loops), suggesting this locus acts as a regulatory hub. However, we observed a similar composition in cell types for four other genes: *DNAJC27*, *DNAJC27-AS1* (both previously implicated in obesity and/or diabetes traits^40^), *AC013267.1*, and *SNORD14* (*RF00016*). *ITSN2*, *NCOA1*, and *EFR3B* were three genes within this locus that were only implicated in immune cell types. *NCOA1* encodes a prominent meta-inflammation factor^41^ known to reduce adipogenesis and shift the energy balance between white and brown fat, and its absence known to induce obesity^42^.

*CALCR* was the most frequently implicated gene at its locus, supported by 20 cell types across all systems. While within the *BDNF* locus, *METTL15* and *KIF18A* – two non-cell-type-specific genes - plus some lncRNA genes, were contacted by childhood obesity-associated proxies within the same multiple cell types as *BNDF*, again suggesting the presence of a regulatory hub.

At the *FAIM2* locus on chr 12q13.12, we observed known genes associated with obesity, eating patterns, and diabetes-related traits, including *ASIC1*, *AQP2*, *AQP5*, *AQP6, RACGAP1*, and *AC025154.2 (AQP5-AS1)* along with *FAIM2* **(Table 1)**. These genes were harbored within cREs of astrocytes, neural progenitors, hypothalamic neurons, and multiple metabolic cell types. Plasmacytoid and CD1c+ conventional dendritic cells were the only two immune cell types that harbored such proxies within their cREs, implicating *ASIC1*, *PRPF40B*, *RPL35AP28*, *TMBIM6*, *and LSM6P2* at the *FAIM2* locus.

The independent *ADCY9* and *FTO* loci are both located on chromosome 16. Genes at the *ADCY9* locus were only implicated in a subset of immune cell types. Interestingly, genes at the *FTO* locus were only implicated in Hi-C datasets (as opposed to Capture C), including 6 metabolic cell types and astrocytes. Most genes at the *FTO* locus were implicated in skeletal myotubes, differentiated osteoblasts, and astrocytes, namely *FTO* and *IRX3*; while *IRX5, CRNDE*, and *AC106738.1* were also implicated in adipocytes and hepatocytes.

### The most implicated cell types by two sets of analyses

EndoC-BH1 and myotubes are the two cell types in which we implicated the most effector genes, with 38 and 42, respectively – **Fig. 3** side panel. This phenomenon is likely proportional in the case of myotubes, given the large number of cREs identified by overlapped Hi-C contact data and ATAC-seq open regions (**Fig. 1A**), but not for EndoC-BH1. Albeit harboring an average number of cREs compared to other cell types, EndoC-BH1 cells were consistently among the top-ranked heritability estimates for the childhood obesity variants resulting from the EGG consortium GWAS (**Fig. 1**) and harbored a significant number of implicated genes by the mapping of proxies. Interestingly, the pancreatic alpha cell type – shown above to be the most significant for heritability estimate by LDSC – revealed only 6 implicated genes contacted by the defined proxies, namely *BDNF* and five lncRNA genes.

### Pathway analysis

Of the 111 implicated genes in total, PubMed query revealed functional studies for 66 genes. The remaining were principally lncRNA and miRNA genes with currently undefined functions (**Table 1**). To investigate how our implicated genes could confer obesity risk, we performed several pathway analyses keeping them either separated for each cell type or pooling into the respective metabolic, neural, or immune system sets. **eFig. 7** shows simple Gene Ontology (GO) biological process terms enrichment results.

Leveraging the availability of our expression data generated via RNA-seq (available for 46 of 57 cell types), we performed pathway analysis. Given that our gene sets from the variant-to-gene process was stringently mapped, the sparse enrichment from normal direct analyses is not ideal for exploring obesity genetic etiology. Thus, we incorporated two methods from the *pathfindR* package^43^ and our customized *SPIA* (details in **eMethods**). The result of 60 enriched KEGG terms is shown in **eFig. 8** (**eTable 4**), with 13 genes in 14 cell types for *pathfindR* and 39 enriched KEGG terms shown in **eFig. 9** (**eTable 5**), with 10 genes in 42 cell types for customized *SPIA*. There were 20 overlapping pathways between the two approaches (yellow rows in **eTable 4&5**) including many signaling pathways such as the GnRH (hsa04912), cAMP (hsa04024), HIF-1 (hsa04066), Glucagon (hsa04922), Relaxin (hsa04926), Apelin (hsa04371), and Phospholipase D (hsa04072) signaling pathways. They were all driven by one or more of these 5 genes: *ADCY3*, *ADCY9*, *CREBBP*, *MMP2,* and *NCOA1*. Interestingly, we observed the involvement of natural killer cells in nearly all the enriched KEGG terms from *pathfindR* due to the high expression of the two adenylyl cyclase encoded genes, *ADCY3* and *ADCY9*, along with *CREBBP*. The *SPIA* approach disregarded the aquaporin genes (given they appear so frequently in so many pathways that involve cellular channels) but highlighted the central role of *BDNF* which single-handedly drove four signaling pathways: the Ras, Neurotrophin, PI3K-Akt, and MAPK signaling pathways. This also revealed the role of *TRAP1* in neurodegeneration.

These two approaches did not discount the role of *FAIM2* and *CALCR*. However, their absence was mainly due to the content of the current KEGG database. On the other hand, these approaches accentuated the role of the *MMP2* gene at the *FTO* locus in skeletal myotubes, given its consistency within the GnRH signaling pathway (**eFig. 10**), which is in line with previous studies linking its expression with obesity^44–46^.

### Supportive evidence by colocalization of target effector genes with eQTLs

The GTEx consortium has characterized thousands of eQTLs, albeit in heterogeneous bulk tissues^47^. To assess how many observed gene-SNP pairs agreed with our physical variant-to-gene mapping approach in our multiple separate cellular settings, we performed colocalization analysis using ColocQuiaL^38^.

282 genes were reported to be associated with the variants within 13 loci from our variant-to-genes analysis. We found 114 colocalizations for ten of our loci that had high conditioned posterior probabilities (cond.PP.H4.abf ≥ 0.8), involving 44 genes and 41 tissues among the eQTLs. We extracted the posterior probabilities for each SNP within each colocalization and selected the 95% credible set as the likely causal variants (complete list in **eTable 6**). Despite sensitivity differences and varying cellular settings, when compared with our variant-to-gene mapping results, colocalization analysis yielded consistent identification for 21 pairs of SNP-gene interactions when considering the analyses across all our cell types, composed of 20 SNPs and 7 genes. Details of these SNP-gene pairs are shown in **Fig. 4A** and **B**.

Of these 20 SNPs, 15 were at the *ADCY3* locus, in LD with sentinel variant rs4077678, and all implicated *ADCY3* as the effector gene in 29 cell types – 15 metabolic, 6 immune, 4 neural cell types and 4 independent cell lines (**Fig. 4C**). Indeed, missense mutations have been previously reported for this gene in the context of obesity^48,49^ while another member of this gene family, *ADCY5*, has also been extensively implicated in metabolic traits^50^.

### Predicting transcription factors (TFs) binding disruption at implicated genes contributing to obesity risk

TFs regulate gene expression by binding to DNA motifs at enhancers and silencers, where any disruption by a SNP can potentially cause dysregulation of a target gene. Thus, we used *motifbreakR* (R package) to predict such possible events at the loci identified by our variant-to-gene mapping. Each variant was predicted to disrupt the binding of several different TFs, thus requiring further literature cross-examination to select the most probable effects. For example, rs7132908 (consistently contacting *FAIM2* in 25 cell types) was predicted to disrupt the binding of 12 different transcription factors. Among them, SREBF1 (**eFig. 11A**) was the only TF that concurred with evidence that it regulates *AQP2* and *FAIM2* at the same enhancer^51^. The full prediction list can be found in **eTable 7**.

To narrow down the list of putative TF binding sites at each variant position, we leveraged the ATAC-seq footprint analysis using the RGT suite^52^. The final set of Motif-Predicted Binding Sites (MPBS) within each cell type ATAC-seq footprints was used to overlap with the genomic locations of the OCRs, and then overlapped with our obesity variants, resulting in annotated 29 variants. Mosaic plot in **eFig. 11B** shows the number and proportions of variants predicted by *motifbreakR* and/or overlapped with MPBS. Insignificant *P*-value from Fisher’s exact test indicated the independence of the two analyses. Only seven variants were found within the cREs for the same TF motifs predicted to be disrupted by *motifbreakR* (**eFig. 11C**). **eFig. 12** outlines the seven variants that *motifbreakR* and ATAC-seq footprint analysis agreed on the TF bindings they might disrupt.

## DISCUSSION

Given the challenge of uncovering the underlying molecular mechanisms driving such a multifactorial disease as obesity, our approach leveraging GWAS summary statistics, RNA-seq, ATAC-seq, and promoter Capture C / Hi-C offers new insights. This is particularly true as it is becoming increasingly evident that multiple effector genes can operate in a temporal fashion at a given locus depending on cell state, including at the *FTO* locus^53^. Our approach offers an opportunity to implicate relevant cis-regulatory regions across different cell types contributing to the genetic etiology of the disease. By assigning GWAS signals to candidate causal variants and corresponding putative effector genes via open chromatin and chromatin contact information, we enhanced the fine-mapping process with an experimental genomic perspective to yield new insights into the biological pathways influencing childhood obesity.

LD score regression is a valuable method that estimates the relationship between linkage disequilibrium score and the summary statistics of GWAS SNPs to quantify the separate contributions of polygenic effects and various confounding factors that produce SNP-based heritability of disease. The general positive heritability enrichment across our open chromatin features spanning multiple cell types (**Fig. 1B.a**) reinforces the notion that obesity etiology involves many systems in our body.

While obesity has long been known to be a risk factor for pancreatitis and pancreatic cancer, the significant enrichment of pancreatic alpha and beta cell related 3D genomic features for childhood obesity GWAS signals demonstrates the bidirectional relationship between obesity and the pancreas; indeed, it is well established that insulin has obesogenic properties. Moreover, the comorbidity of obesity and diabetes (either causal or a result of the overlap between SNPs associated with these two diseases) is tangible. When focusing on genetic annotation of the cREs only, the association with obesity became more diverse across cell types, especially in metabolic cells. Interestingly, the lack of enrichment (only 8 of 57 cell types yielded no degree of enrichment) of obesity SNPs heritability in open gene promoters (**Fig. 1B.b**) reveals that cRE regions harboring obesity SNPs are more involved in gene regulation than disruption, and therefore potentially contributing more weight to the manifestation of the disease.

Of course, we should factor in the effective sample sizes of the GWAS efforts that are wide-ranging (2,000-24,000 – given that the N for each variant is different within a single dataset, thus contributing to the weights and *P*-value of each SNP when the algorithm calculates the genome-wide heritability), which could result in noise and negative enrichment observed in the analysis – a methodology limitation of partial linkage regression that has been extensively discussed in the field^54^. Thus, it is crucial to interpret the enrichment (or lack thereof) of disease variants in a certain cellular setting with an *ad hoc* biological context.

From mapping the common proxies of 19 independent sentinel SNPs that were genome-wide significantly associated with childhood obesity to putative effector genes through chromatin contacting cREs, one striking finding was the several potential “hubs” of putatively core effector genes, whose occurrence spread across three human physiological systems. With the data available from so many cell types, our approach connected new candidate causal variants to known obesity-related genes and new implications of cell modality for previously known associations.

A potential application of this association could be to fine-tune the effect of a drug toward controlling appetite. An example of bringing new aspects to the old is for the signal within the *FTO* locus that contacted *IRX3* and *IRX5*: previous studies have suggested these obesogenic effects operate in adipocytes^55^, brain^56^, or pancreas^57^; here we confirmed this association in adipocytes and uncover the presence of distal chromatin contacts in myotubes for the first time.

Besides the above-mentioned genes with known associations with obesity, we discovered newly implicated genes. For example, the *LRRIQ3* gene at the *TNNI3K* locus had its open promoter contacted by two SNPs, rs1040070 and rs10493544, in NTERA2 cells only. The published studies^58,59^ that associated *LRRIQ3* with major depressive disorder and opioid usage acknowledged the overlapping promoter of this gene, albeit in the opposite direction, with a run-through transcript of *FPGT*-*TNNI3K* – previously shown to be associated with BMI in European^60^ and Korean populations^61^.

It is apparent that not all the implicated genes we report would contribute equally to the susceptibility of obesity pathogenesis. Each locus comprises genes whose functions are obviously related to obesity or similar traits like BMI, fat weight, etc., while other genes are not so directly obvious in their relation to these traits.

It is encouraging that for implicated genes within these multi-cell-type loci across different physiological systems we could find previous associations to the corresponding cell types or systems. Examples are the two aforementioned genes at the *TMEM18* locus (*SH3YL1* and *ACP1*)^62–66^ with the broad spectrum of their functions, *HEPACAM2* implicated in the NCIH716 cell line at the *CALCR* locus^67,68^, and *LRRIQ3* in the NTERA2 cell line at the*TNNI3K* locus^69^.

Chronic inflammation is an essential characteristic of obesity pathogenesis. Adipose tissue-resident immune cells have been observed, leading to an increased focus in recent years on their potential contribution to metabolic dysfunction. On the other hand, neurological or psychological conditions, such as stress, induce the secretion of both glucocorticoids (increase motivation for food) and insulin (promotes food intake and obesity). Pleasure feeding then reduces activity in the stress-response network, reinforcing the feeding habit. It has been shown that voluntary behaviors, stimulated by external or internal stressors or pleasurable feelings, memories, and habits, can override the basic homeostatic controls of energy balance^70^. The potential link between the immune system and metabolic disease, and moreover, through the neural system, was tangible in our findings.

Two of the three SNPs which ranked the third most consistent in our variant-to-gene mapping (**Fig. 2C**) – rs35796073 and rs35142762 – contacted the *ALKAL2* promoter (supported by GTEx evidence to colocalize with *ALKAL2* expression). The anaplastic lymphoma kinase (encoded by *ALK* gene) is a receptor tyrosine kinase, belongs to the insulin receptor family, and has been reported to promote nerve cell growth and differentiation^71,72^. Despite *ALKAL2* (ALK and LTK ligand 2) being studied principally in the context of immunity, a recent study using the EGCUT biobank GWAS identified *ALK* as a candidate thinness gene and genetic deletion showed that its expression in hypothalamic neurons acts as a negative regulator in controlling energy expenditure via sympathetic control of adipose tissue lipolysis^73^. *ALKAL2 –* encoding a high-affinity agonist of *ALK*/*LTK* receptors – which has been reported to enhance expression in response to inflammatory pain in nociceptors^74,75^ - has been recently implicated as a novel candidate gene for childhood BMI by transcriptome-wide association study^76^, and achieved genome-wide significance in a GWAS study contrasting persistent healthy thinness with severe early-onset obesity using the STILTS and SCOOP cohorts^77^. The finding that overexpression of *ALKAL2* could potentiate neuroblastoma progression in the absence of ALK mutation^78^ echoes the relationship between *ADCY3* and *MC4R*^79^, where a peripheral gene, *ADCY3*, can regulate/impair the function of a core gene, i.e. *MC4R*, within the energy-regulating melanocortin signaling pathway^80^.

Our approach implicates putative target genes based on a mechanism of regulation for these variants to alter gene expression – through regulator TF(s) that bind to these contact sites. A potential limitation of the predictions from *motifbreakR* and matching TF motifs to ATAC-seq footprint by the RGT toolkit is that they were both based on the position probability matrixes of Jaspar and Hocomoco, which come from public motif databases. The ATAC-seq footprint analysis also carries sequence bias that can lead to false positive discovery. Thus, our attempt to call such regulators by predicting TF binding disruption can only serve as nominations – but warrant further functional follow up.

Another limitation of this work is the diversity in data quality among different samples, since different datasets were sampled and collected at different time points, from different patients, using different protocols, with libraries sequenced at different depths and qualities, and initially preprocessed with different pipelines and parameters. Thus, it is crucial to keep in mind that the discrepancy in data points might have resulted from variations in data quality. Importantly, any association discovered must be validated functionally before effector genes of the genetic variants can be leveraged to develop new therapies. Their putative function(s) must be characterized, together with the mechanism whereby the given variant’s alleles differentially affect the expression of the targeted genes. The next step is to explore how the target genes affect the trait of interest more directly.

Our results have provided a set of leads for future exploratory experiments in specific cellular settings in order to further expand our knowledge of childhood obesity genomics and hence equip us with more effective means to overcome the burden of this systematic disease.

## CONCLUSION

Our approach of combining RNA-seq, ATAC-seq, and promoter Capture C/Hi-C datasets with GWAS summary statistics offers a systemic view of the multi-cellular nature of childhood obesity, shedding light on potential regulatory regions and effector genes. By leveraging physical properties, such as open chromatin status and chromatin contacts, we enhanced the fine-mapping process and gained new insights into the biological pathways influencing the disease. Although further functional validation is required, our findings provide valuable leads together with their cellular contexts for future research and the development of more effective strategies to address the burden of childhood obesity.

## Supporting information

eTables1-7

eMethods, eFigures and legends

## Data Availability

All data produced in the present study are available upon reasonable request to the authors

## ACKNOWLEDGMENTS

This work was supported by National Institutes of Health awards R01 HD056465, R01 DK122586 and UM1 DK126194, and the Daniel B. Burke Endowed Chair for Diabetes Research.

Given the use of de-identified datasets and biospecimens was not considered human subjects research, ethical oversight was waived by the Institutional Review Board of the Children’s Hospital of Philadelphia

