## Supplementary material for "3D genomic features across >50 diverse cell types reveal insights into the genomic architecture of childhood obesity": eMethods, eFigures and legends

***ATAC-seq preprocessing and peaks calling:*** The detailed configurations and technical details for each data set are provided in the original studies. In brief, open chromatin regions were called using ENCODE ATAC-seq pipeline as previously described in each study the published data provided. In brief, reads were aligned to hg19 or hg38 genome using bowtie2, duplicates were removed, alignments from all replicates were pooled, narrow peaks were called using MACS2. A region was considered open if it overlaped at least 1bp with ATAC-seq peak. We lifted all coordinates from hg19 to hg38 to ensure consistency between datasets.

***ATAC-seq transcription factor footprint analysis with RGT toolkit:*** Bam files of mapped reads from replicates and samples were merged for each cell type. The merged bam files were then used for footprinting by RGT-HINT with parameters: *--atac-seq, –paired-end, –organism=hg38*. If bam files were generated on hg19, we performed lift-over using CrossMap.py *bam* and hg19ToHg38.over.chain.gz file. We then used RGT-MOTIFANALYSIS *matching* to scan each footprint for possible transcription binding sites from HOCOMOCO and JASPAR databases for human only with parameter --filter "species:sapiens;database:hocomoco,jaspar_vertebrates". Parameter *–rand-proportion 10* was used to generate random putative binding sites with sizes ten times larger than the input footprints. After performing motif matching, we evaluated which transcription factors were more likely to occur in those footprints than in background regions (generated by the previous command) using RGT-MOTIFANALYSIS *enrichment* with the same filtered databases and default parameters. Output included all the Motif-Predicted Binding Sites (MPBS) that occurred within the identified footprints in each cell type. We overlapped these sites with the loci of our obesity variants.

***RNA-seq preprocessing and expression profiling:*** The detailed configurations, steps, and technical details for each data set are provided in the original studies. In brief, read fragments from fastq files were mapped to genome assembly hg19 or hg38 using STAR, independently for each replicate and condition. We used GENCODE annotation files for feature annotation and htseq-count for raw read count calculation at each feature. Read counts were transformed into TPM (transcript per million) and normalized internally between replicates/conditions in each individual study. For comparative measurements, we transformed all the expression values into 0-100 scale.

***Promoter Capture-C pre-processing and interaction calling:*** in brief, paired-end reads were pre-processed using HICUP pipeline^1^ with bowtie2 as aligner and hg19 for reference genome. Significant promoters’ interactions were called using unique read pairs from all baits promoter in the reference by CHICAGO^2^ pipeline. In addition to analysis of individual fragments (1frag), we also binned four fragments to improve long-distance sensitivity in interactions calling^3^. Interactions with CHICAGO score > 5 in either 1-fragment or 4-fragment resolution were considered significant. These interactions were output as *ibed* format (similar to BEDPE format) in which each line represents one physical contact between fragments. Interactions from both resolutions were merged and their genomic coordinates were lifted from hg19 to hg38.

***Hi-C pre-processing and interaction calling:*** We follow the pipeline as a recent study described^4^. Paired-end reads from each replicate were pre-processed using the HICUP pipeline v0.7.4^1^, aligned by bowtie2 with hg38 as the reference genome. The alignments files were parsed to pairtools v0.3.0 to process and pairix v0.3.7 to index and compress, then converted to Hi-C matrix binary format .*cool* by cooler v0.8.11 at multiple resolutions (500bp, 1, 2, 4, 10, 40, 500kbp and 1Mbp) and normalized with ICE method^5^. The matrices from different replicates were merged at each resolution using cooler. Mustache v1.0.1^6^ and Fit-Hi-C2 v2.0.7^7^ were used to call significant intra-chromosomal interaction loops from merged replicates matrices at three resolutions 1kb, 2kb, and 4kb, with significance threshold at q-value < 0.1 and FDR < 1 × 10^−6^, respectively. The identified interaction loops were merged between both tools at each resolution. Lastly, interaction loops from all three resolutions were merged with preference for smaller resolution if overlapped.

***Differential analysis and clustering of correlated genes:*** Normalized transcripts per million (TPM) of all measured genes in 46 of 57 cell types was used to perform differential analysis using ***DEseq2*** package^8^, where cell type and system (immune, metabolic, neural and other) were used as variables for the modeling contrast. Because many of the genes we gathered from the variant-to-gene mapping were lowly expressed in corresponding cell types or others, causing relatively high levels of variability, we used *apeglm* method for effect size (logarithmic fold change estimates) shrinkage^9^ to alleviate this phenomenon during the genes ranking. Weighted correlation network analysis (***WGCNA***) package^10^ was used to cluster genes from the variant-to-genes mapping process. WGCNA network construction power was chosen based on the analysis of scale-free topology for soft-thresholding. *blockwiseModules* with a power of 10 was used to create the correlation network and cluster genes into modules of similarly expressed genes.

***Pathways enrichment analyses:*** We performed three analyses on the set of genes from the variant-to-genes mapping process:

***Gene set over-representation analysis (ORA)*** was performed using ***clusterProfiler*** package^11^ to identify GO biology process terms (org.Hs.eg.db databese) enriched within our genes set in each cell type. A relaxed P-value cutoff was set at 0.1, and the minimum including genes was set at 2 to ensure the capture of all possible enriched terms. An adjusted P-value of 0.05 was later used to filter the significant terms.

***Active-subnetwork-oriented gene set enrichment analysis***: We used the ***pathfindR*** package^12^ to identify active subnetworks in protein-protein interaction networks from Biogrid, KEGG, STRING, GeneMania, and IntAct databases, using the list of genes from the variant-to-genes mapping process. Then we provided the statistic from the differential expression analysis for ***pathfindR*** to perform enrichment analyses on the identified subnetworks, discovering enriched KEGG pathways.

***Customized Signaling Pathway Impact Analysis (SPIA)***: The original method, proposed by Tarca in 2009 in ***SPIA*** package^13^, incorporates ORA with the adjacency matrix to measure the importance of genes within each pathway – genes that are connected to more other genes are likely more important than the downstream end-point genes. This pathway-topology approach measures the actual perturbation on a given pathway under a given condition and given differential effect size. We added more metrics to improve this method: a) the score of gene impact among networks, b) the neighborhood of genes that measure the importance of a gene based on its downstream effects, c) the betweenness puts weight on the genes that act as a gateway for the network flow. The combination of metrics produces two ways of evidence – perturbance and enrichment – for each pathway similar to *SPIA*. Normal inverse cumulative distribution function was used to combine the P-values of this evidence, then Bonferroni and FDR correction was applied.

***Prediction of variant’s effect on transcription factor binding:*** Genomic positions (0-based coordinates) and allele alternatives of each proxy (from ***SNPlocs.Hsapiens.dbSNP155.GRCh38*** package with matching reference sequence from ***BSgenome*** package) were used to scan all position frequency matrix databases (from ***MotifDb*** package) for potential transcription factor binding disruptive effects. The motifbreakR function from ***motifBreakR*** package was used, with *filterp=TRUE* and setting a p-value *threshold=0.0005*, information content methods *method='ic'* with even background probabilities of the four nucleotides *bkg = c(A=0.25, C=0.25, G=0.25, T=0.25)* and *BPPARAM = BiocParallel::SerialParam()* to allow serial evaluation.

**eFigures and legends**

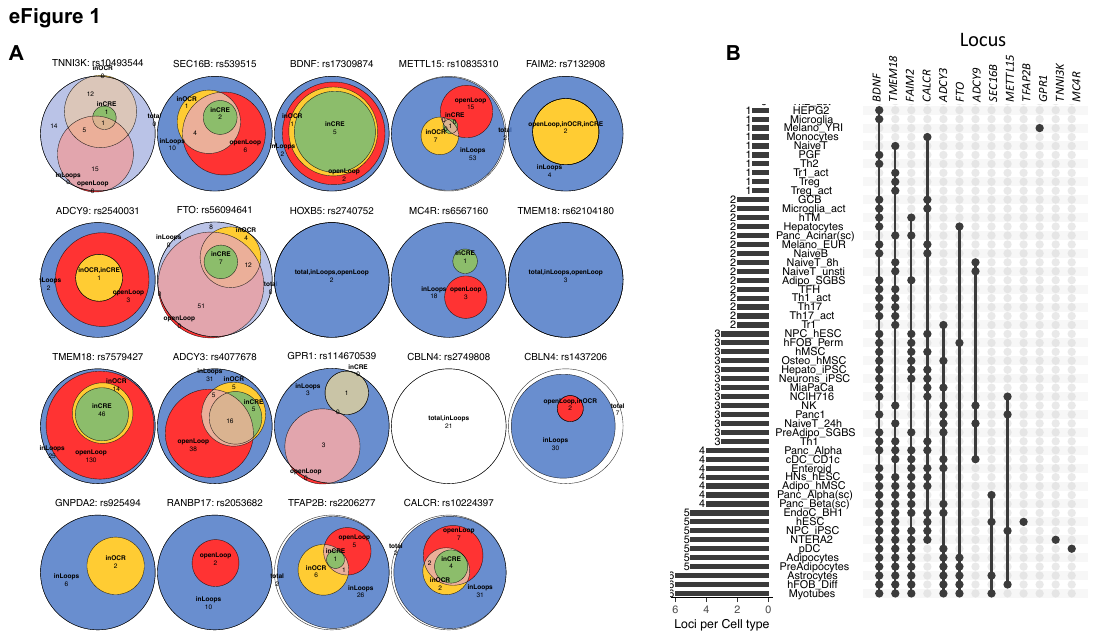

**eFigure 1: Mapping statistics of the childhood obesity-associated loci**

A. Venn diagrams show intersections and the number of proxies within each locus were mapped in different scenarios as illustrated in **Fig1A** - colors of the areas corresponding to different subset of OCRs, the white areas show proxies that unmapped to any OCR.
B. The upSet plot outlines in which cell type(s) each locus appeared.

The 46 variants at the *TMEM18* locus appeared in 31 cell types and implicated 16 genes, while only 5 variants of *BDNF* locus appeared in 42 cell types and implicated 23 genes, or 2 variants of *FAIM2* locus with 26 cell types and 15 implicated genes. Seven variants of the *FTO* locus though appeared in only 7 cell types, but targeted promoters of up to 18 genes. At five loci *ADCY9*, *GPR1*, *MC4R*, *METTL15,* and *TFAP2B*, only one single proxy – but not the lead variant in each case – was mapped. Of these, the variant from *METTL15* locus only implicated the *BDNF* gene but appeared in four different cell types. In contrast, the variant from *ADCY9* locus appeared in five different cell types and implicated up to 10 different genes.

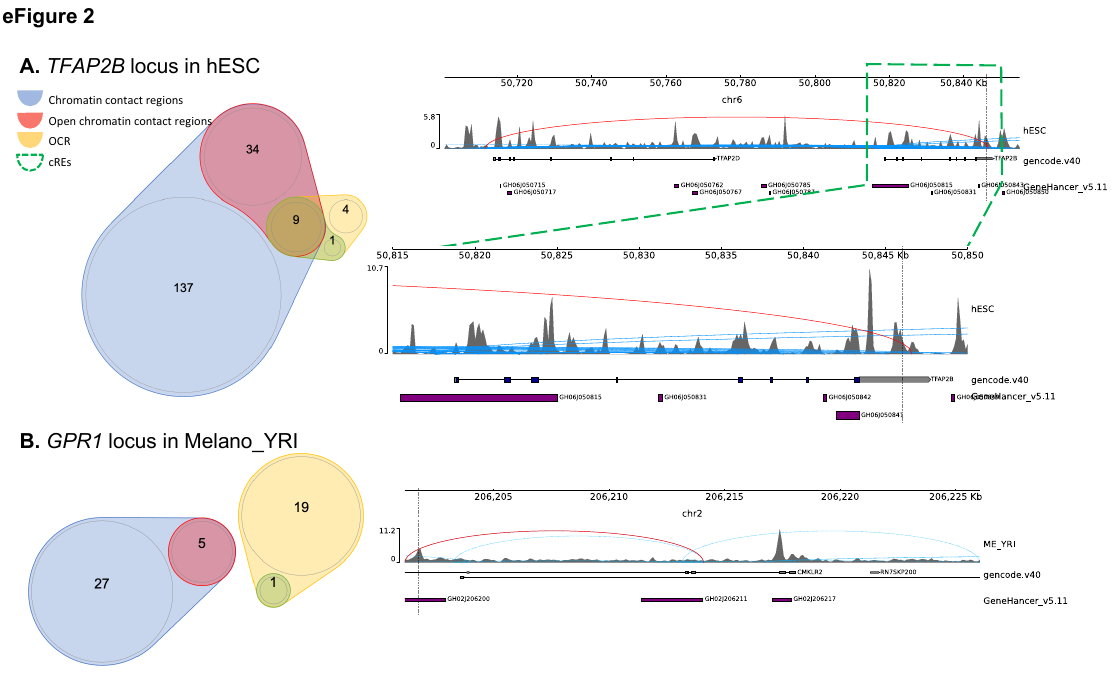

**eFigure 2: Examples of 2 variants mapped to only one gene promoter**

A. At *TFAP2B* locus on chromosome 6, rs62405437 was found only in a human embryonic stem cell line, within the body of transcription factor AP-2 beta (*TFAP2B)* contacting the promoter of *TFAP2D* (red arc). Interestingly, there were many contacts identified by our Capture-C assay between these two members of the transcription factor AP-2 family (blue arcs), and many of our proxies were located within these loop ends, albeit outside any defined open chromatin region – red regions in illustration **Fig1A**
B. At the *GPR1* locus on chromosome 2, rs115319174 was identified only within Nigerian melanocytes cREs, positioned in an open region within the body of the *CMKLR2* gene and contacting (red arc) the promoter of *GPR1* – a gene for G protein-coupled receptors known to increase its expression in obese phenotypes^14^. Blue arcs represented other chromatin contacts identified within this window.

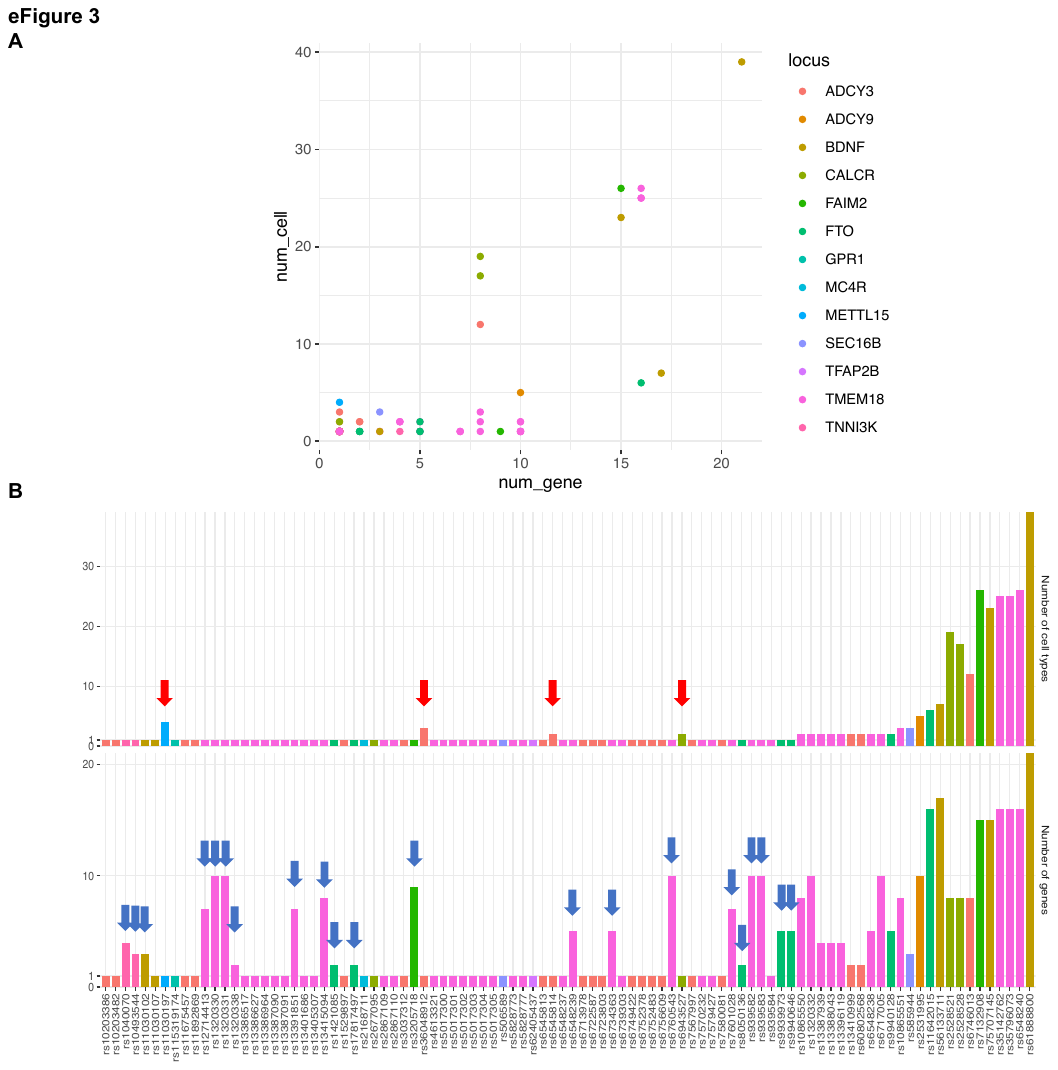

**eFigure 3: The number of genes and cell types each proxy mapped into our cREs**

A. Dot-plot with each variant colored by their locus
B. Bar-plot layout. Some variants found in multiple cell types were more selective with respect to their target genes (**red arrow**) – such as rs11030197 at the *METTL15* locus – or conversely more selective in given cell types but implicated multiple genes (**blue arrow**) – such as 21 other variants from the 66 mentioned earlier that appeared in only one cell type but contacted multiple genes within that cellular setting. These 21 variants included one from *BDNF* locus, one from *FAIM2* locus, two from *TNNI3K*, five from *FTO,* and twelve from the *TMEM18* locus.

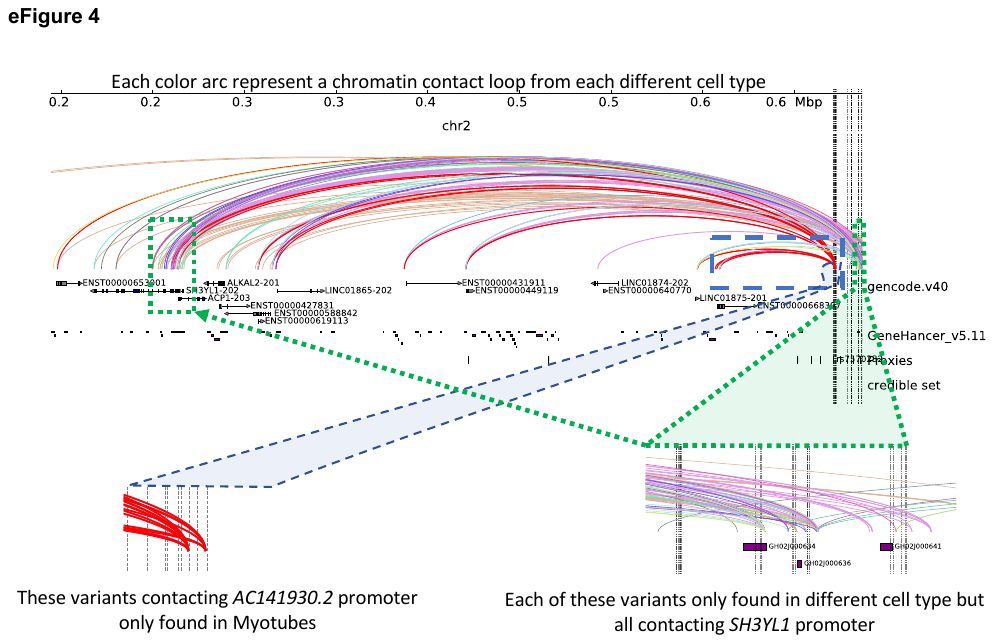

**eFigure 4:** **The *TMEM18* locus**

Within the *TMEM18* locus where there were 45 proxies in LD with the sentinel SNP rs7579427 that mapped to the cREs of 31 cell types; 35 of these proxies were found exclusively in only one unique cell type (10 in pre-adipocytes only, 9 in adipocytes only, 8 in EndoC-BH1 only, 2 in hESC-derived hypothalamic neurons only, 2 in natural killer cells only, and 4 in activated regulatory T cells only); however, their target gene promoters were also frequently contacted by other different proxies in other cell types. Example in **blue window**.

**The green window:** The *SH3YL1* open promoter was contacted within 25 cell types, including eleven immune cell types (mostly T-cells), six metabolic cell types (adipocytes, different pancreatic cell types, differentiated osteoblasts), and three neural cell types (primary astrocytes, hESC-derived hypothalamic neurons, and neural progenitors). Similarly, the *ACP1* promoter was contacted in 14 cell types across three systems, except for adipocytes and astrocytes.

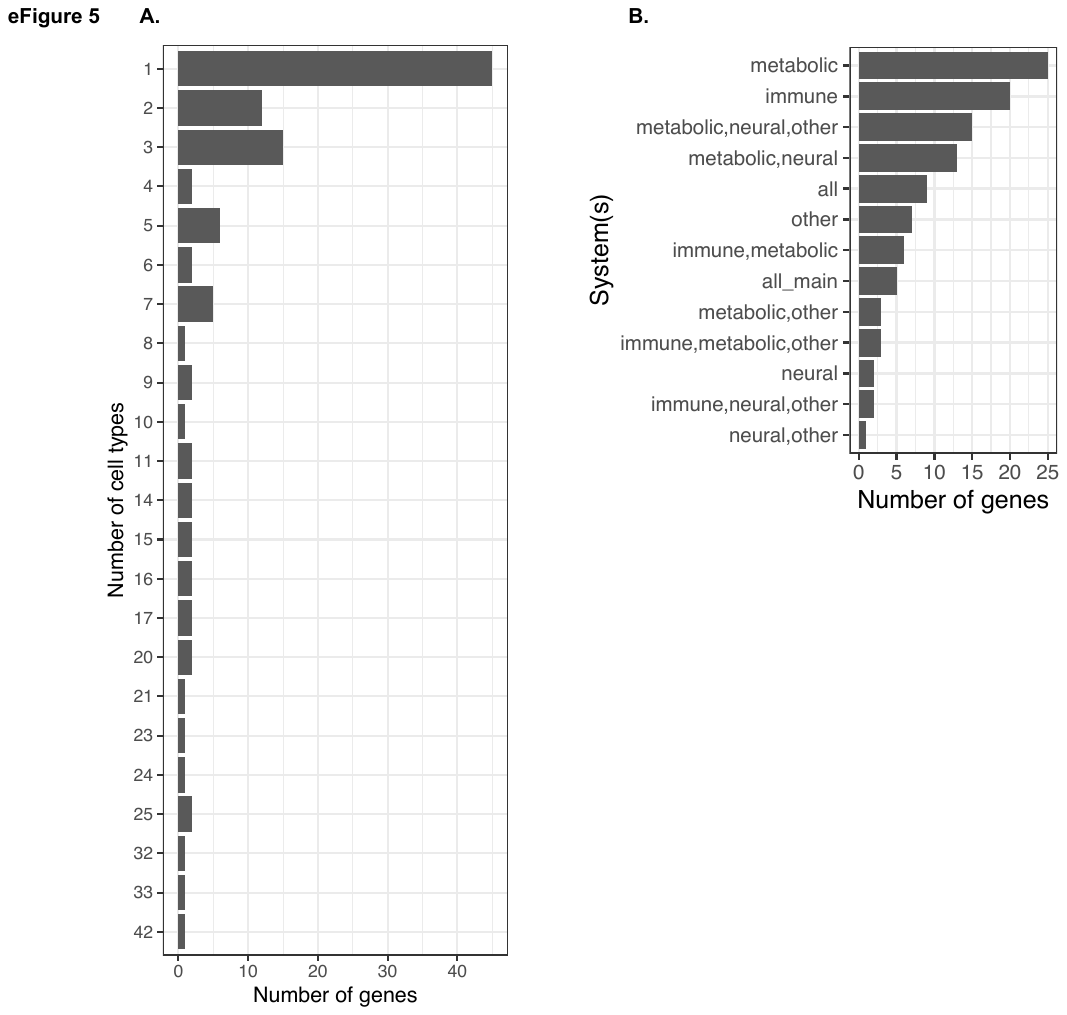

**eFigure 5: The statistics of the 111 implicated genes**

A. Bar-plot shows number of genes implicated in how many cell types.
B. Bar-plot shows number of genes implicated in cell types of each combination of ***metabolic***, ***immune***, ***neural*** system and ***other*** cell lines groups.

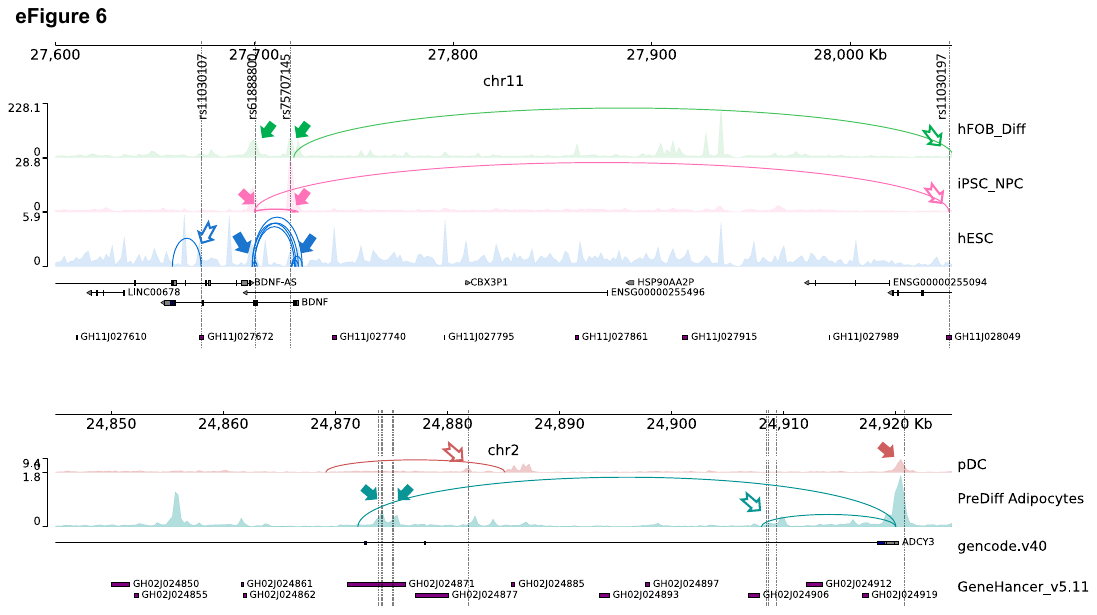

**eFigure 6: Double-implicated genes**

Genomic views of two loci where genes promoters both harbor childhood obesity variants (solid arrows é) and were contacted by OCRs harboring variants though chromatin contacts (hollow arrows ñ), hence “double-implicated”.

Top panel: *BDNF* locus where *BDNF* gene promoters harbor two variants rs61888800 and rs75707145 that overlapped with OCRs of 3 cell types. In differentiated hFOB cell and iPSC-derived neural progenitor cells, different *BDNF* promoters were contacted through chromatin loops in different cell types.

Lower panel: ADCY3 locus where ADCY3 gene promoters were double-implicated by 2 variants in plasmacytoid dendritic cells and 8 other variants in pre-differentiated adipocytes.

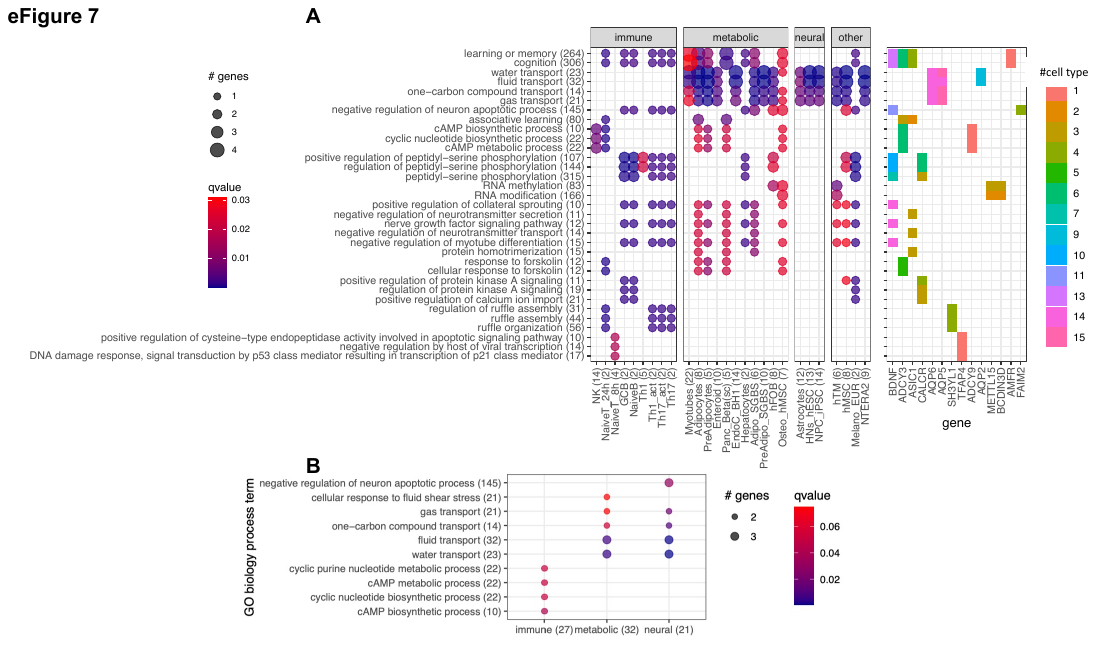

**eFigure 7: Gene Ontology (GO) biological process terms enrichment**

A. Due to the limited number of implicated genes, over-representation analysis for GO biological process terms across the cell types showed only a few main clusters. This result mainly emphasized the frequencies of several genes that appeared in many cell types and participated in many biological processes and pathways – namely *BDNF, FAIM2, CALCR,* Aquaporin genes (*AQP2*, *AQP5*, *AQP6*), *ASIC1,* and the Adenylate cyclase genes (*ADCY3*, *ADCY9*, *BCDIN3D*, *METTL15*).
B. Pooling genes resulting from the 3 systems of metabolic, neural, and immune cell types revealed two clear clusters of processes, segregated by the implicated genes enriched mainly in immune cell types (primarily within the *ADCY3* locus) versus genes enriched on both metabolic and neural cell types. This somewhat reflected the gene clusters in **Figure 3**.

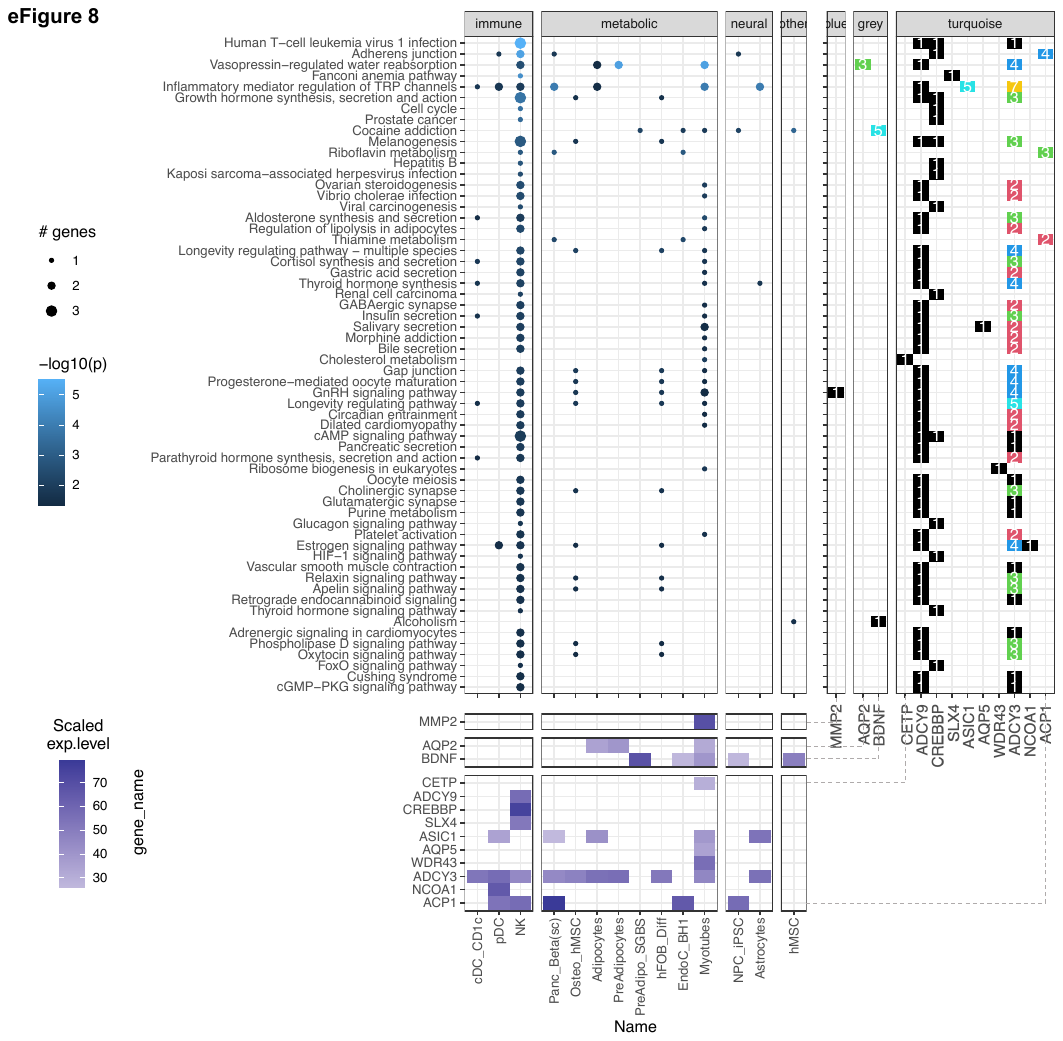

**eFigure 8: The *pathfindR* method**

Focused on “leveraging interaction information from a protein-protein interaction network (PIN) to identify distinct active subnetworks and then perform enrichment analyses on these subnetworks”, thus aiding enriched pathway analyses through the inter-connection between the genes targeted by obesity variants with key genes driving the pathology of the disease. The result with 60 enriched KEGG terms in the main panel (**eTable 4**), shows 13 genes in 14 cell types and their scaled expression levels in the lower panel.

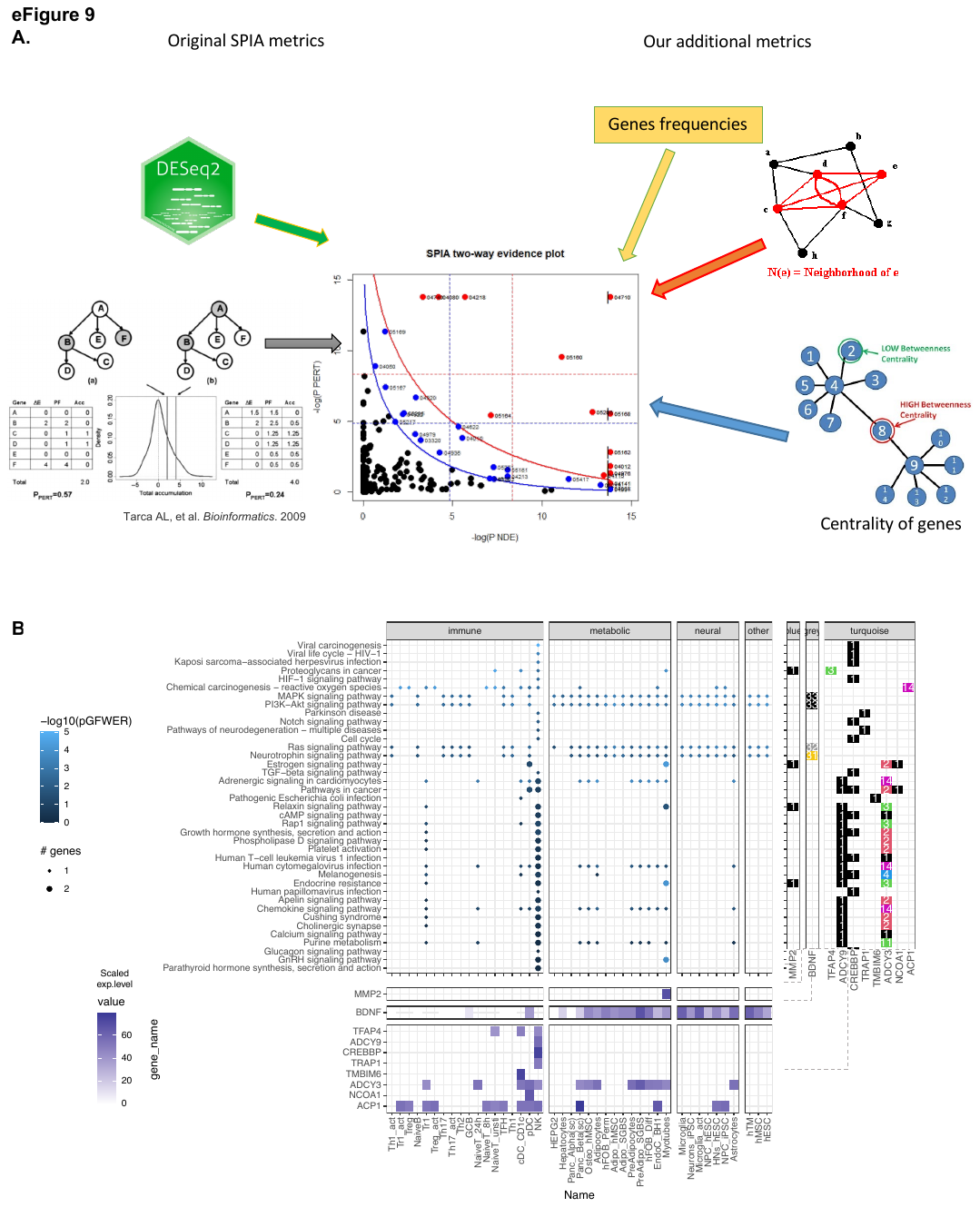

**eFigure 9: modified SPIA analysis**

A. The additional metrics scheme: To extend the analysis beyond the prerequisite protein-protein networks, but still account for how our genes were weighted within the network architecture of the pathways, we devised a modified SPIA analysis with additional metrics
B. After applying the adjusted *P*-value of 0.05 as the filtering threshold, the analysis yielded 39 enriched KEGG terms (**full table at eTable 5**) with only 10 genes, but involved up to 42 cell types.

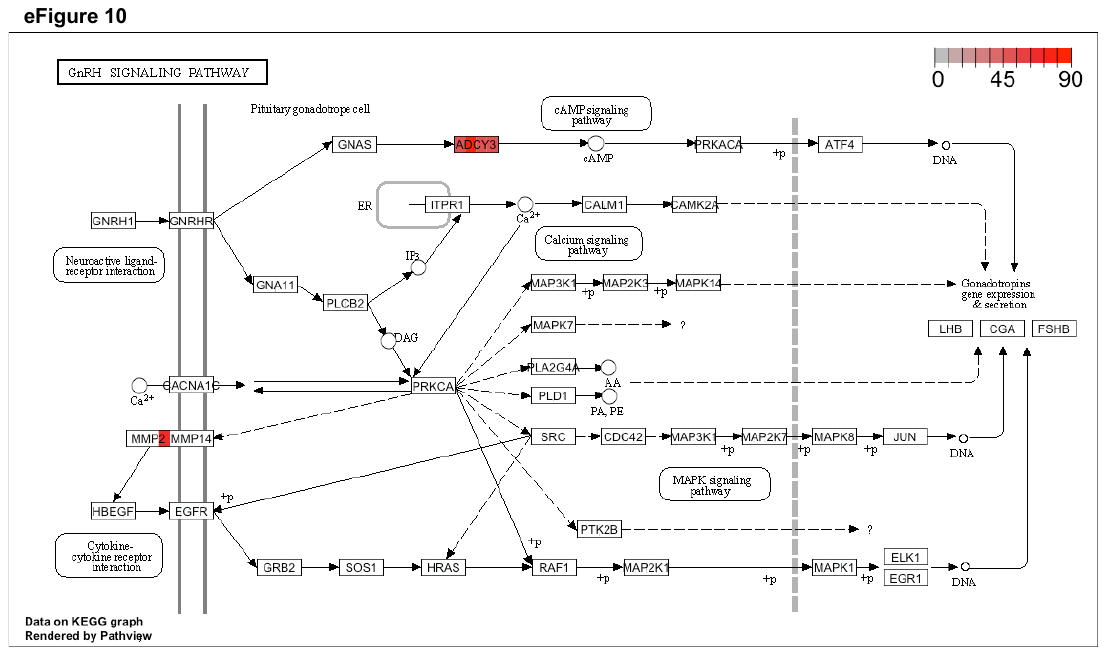

**eFigure 10: The GnRH signaling pathway**

The KEGG graph shows the involvement of *ADCY3* and *MMP2* genes driving the GnRH signaling pathway. The 4 areas in each gene were colored by expression levels of such gene within the corresponding cell types: the hMSC-derived BMP2 differentiated osteoblast, the differentiated osteoblast cell line, the natural killer cell, and the skeletal myotubes.

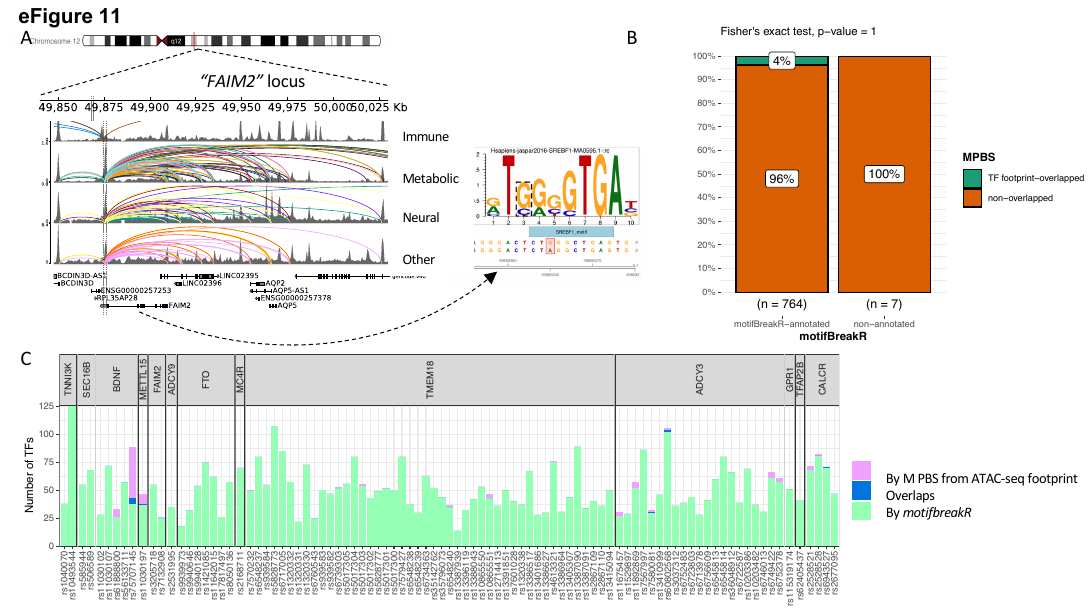

**eFigure 11: *motifbreakR* vs ATAC-seq footprint analysis**

A. Genome view at the *FAIM2* locus where rs7132908 is located and can target many genes through many chromatin contacts, presented by arcs (different colors for different cell types). rs7132908 was predicted by *motifBreakR* to disrupt the TF SREBF1’s binding site, thus potentially altering the expression of its implicated genes.
B. Mosaic plot shows number of variants that were annotated with disrupt-TF-binding affect by motifBreakR, and the proportions that also overlapped with predicted TF footprint from ATAC-seq TF footprint analysis. Fisher exact test was performed and produced P-value = 1.
C. Stacked bar plot for all the variants from variant-to-gene analysis, showing number of transcription factor binding sites each of the variant can disrupt (predicted by *motifbreakR* – green), or simply overlap (analyzed by RGT suite – purple), or both (blue).

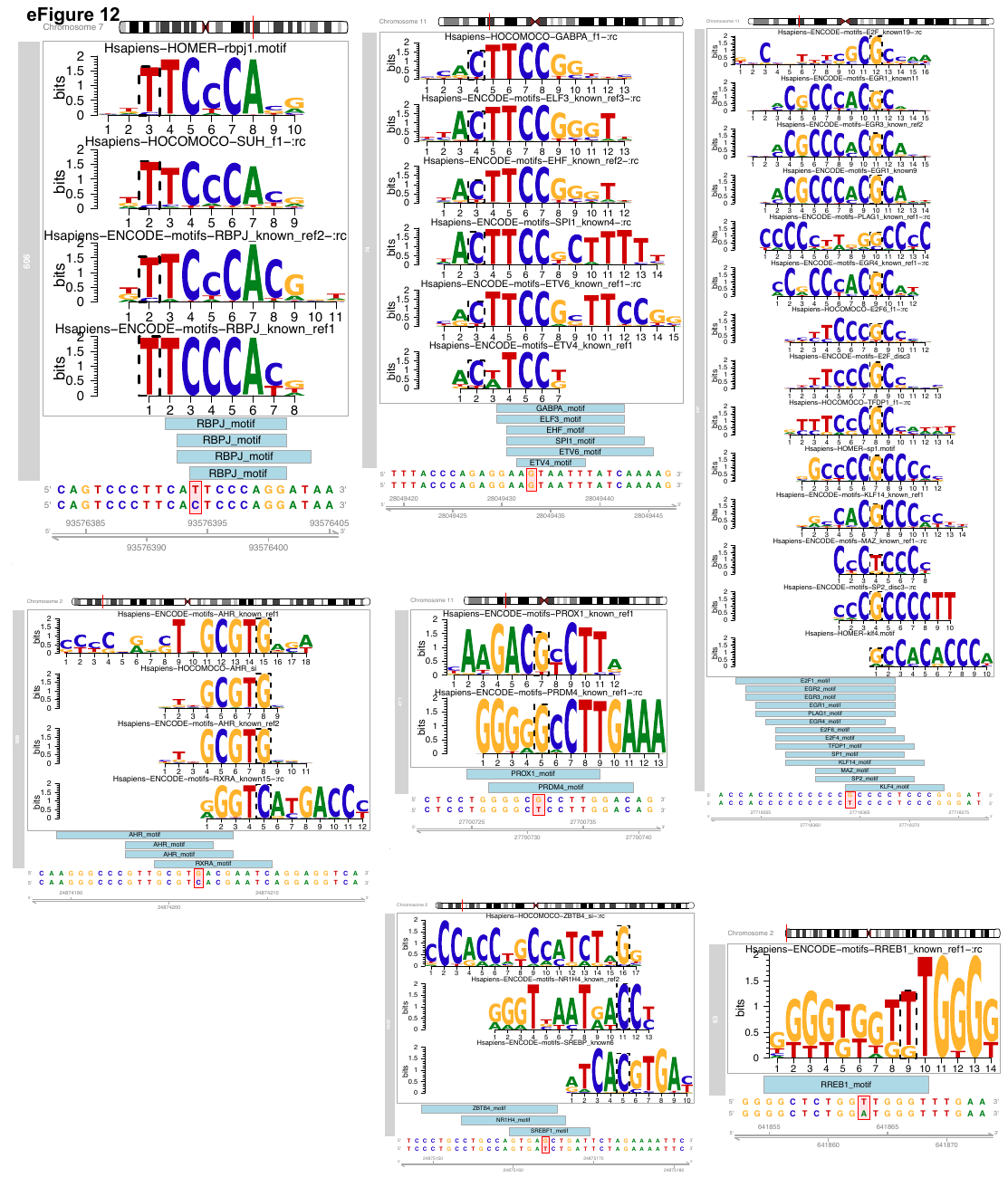

**eFigure 12: Genome view of the 7 variants and the corresponding TF motifs**

A. At the *TMEM18* locus, rs10865551 was predicted to disrupt the binding site of RREB1, in pancreatic alpha and beta cells (from the single-cell data set) by *motifbreakR*, which were supported by cREs-MPBS from ATAC-seq footprints from these cell types; the target genes through chromatin contact were different between these cell types, where rs10865551 contacts the *AC141930.2* promoter in alpha cells, but contacts *SH3YL1* and *ACP1* promoters in beta cells. About eleven thousand bases upstream from that was rs5017303, which was predicted to disrupt binding sites of BC11A in adipocytes but contacts the same gene *AC141930.2* as in pancreatic alpha cells.
B. rs60802568 in *ADCY3* locus was predicted to disrupt bindings of SREBF1 and SREBF2 by *motifbreakR*, which were supported by cREs-MPBS from ATAC-seq footprints from astrocytes, pre-adipocytes, adipocytes and osteocytes derived from mesenchymal stem cells. It was also predicted to disrupt ZBTB4 and NR1H4 with supported cREs-MPBS from ATAC-seq footprints only in osteocytes derived from mesenchymal stem cells.
C. Another variant from *ADCY3* locus, rs7580081 was predicted to disrupt AHR binding in pre-adipocytes and RXRA binding in adipocytes.
D. At the *CALCR* locus, rs6943527 was predicted to disrupt RBPJ binding in naïve T-cells that got activated for 24h.
E. From *METTL15* locus, rs11030197 is one of the few variants that only targets one single gene promoter, namely *BDNF* gene, but is implicated in many cell types. It was predicted to disrupt the binding sites of 6 different TFs in melanocytes and enteroids.
F. The “regulatory-hub” variant rs61888800 of *BDNF* locus was found to contact multiple genes across 41 different cell types and was predicted by *motifbreakR* to disrupt binding sites of many TFs. The overlaps with cREs-MPBS only provided support for disruption of binding of PROX1 and PRDM4 in germinal center B cell-like cell line, naïve B cells, naïve T cells, and the HEPG2 cell line where it contacts via short-ranged chromatin loops with the *BDNF* promoter, and long-ranged chromatin loops with several lncRNA genes: *AC090833.1*, *AC100773.1*, *AC090791.1* and *AC013714.1*.
G. Another variant from *BDNF* locus, rs75707145 is predicted to disrupt at least 14 TFs binding sites, supported by overlaps with cREs-MPBS in pre-adipocytes, pancreatic cells, permissive osteoblasts, EndoC BH1 cell and NCIH716 cell lines.
